# Long-term and serious harms of medical cannabis and cannabinoids for chronic pain: A systematic review of non-randomized studies

**DOI:** 10.1101/2021.05.27.21257921

**Authors:** Dena Zeraatkar, Matthew Adam Cooper, Arnav Agarwal, Robin W. M. Vernooij, Gareth Leung, Kevin Loniewski, Jared E. Dookie, Muhammad Muneeb Ahmed, Brian Younho Hong, Chris J. Hong, Patrick Jiho Hong, Rachel Couban, Thomas Agoritsas, Jason W. Busse

## Abstract

**Objective:** To establish the risk and prevalence of long-term and serious harms of medical cannabis and cannabinoids for chronic pain.

**Design:** Systematic review and meta-analysis.

**Data sources:** MEDLINE, EMBASE, PsycInfo, and the Cochrane Central Register of Controlled Trials (CENTRAL) from inception to April 1, 2020.

**Study selection:** Non-randomized studies reporting on harms of medical cannabis or cannabinoids in people living with chronic pain with ≥4 weeks of follow-up.

**Data extraction and synthesis:** A parallel guideline panel provided input on the design and interpretation of the systematic review, including selection of adverse events for consideration. Two reviewers, working independently and in duplicate, screened the search results, extracted data, and assessed risk of bias. We used random-effects models for all meta-analyses and the GRADE approach to evaluate the certainty of evidence.

**Results:** We identified 39 eligible studies that enrolled 12,143 patients with chronic pain. Very low certainty evidence suggests that adverse events are common (prevalence: 26.0%; 95% CI 13.2 to 41.2) among users of medical cannabis or cannabinoids for chronic pain, particularly any psychiatric adverse events (prevalence: 13.5%; 95% CI 2.6 to 30.6). However, very low certainty evidence indicates serious adverse events, adverse events leading to discontinuation, cognitive adverse events, accidents and injuries, and dependence and withdrawal syndrome are uncommon and typically occur in fewer than one in 20 patients. We compared studies with <24 weeks and ≥ 24 weeks cannabis use and found more adverse events reported among studies with longer follow-up (test of interaction p < 0.01). Palmitoylethanolamide was usually associated with few to no adverse events. We found insufficient evidence addressing the harms of medical cannabis compared to other pain management options, such as opioids.

**Conclusions:** There is very low certainty evidence that adverse events are common among people living with chronic pain who use medical cannabis or cannabinoids, but that few patients experience serious adverse events. Future research should compare long-term and serious harms of medical cannabis with other management options for chronic pain, including opioids.

**Systematic review registration https://osf.io/25bxf**

**What is already known on this topic:**

- Medical cannabis and cannabinoids are increasingly used for the management of chronic pain.
- Clinicians and patients considering medical cannabis or cannabinoids as a treatment option for chronic pain require evidence on benefits and harms, including long-term and serious adverse events to make informed decisions.

**What this study adds:**

- Very low certainty evidence suggests that adverse events are common among people living with chronic pain who use medical cannabis or cannabinoids, including psychiatric adverse events, though serious adverse events, adverse events leading to discontinuation, cognitive adverse events, accidents and injuries, and dependence and withdrawal syndrome are uncommon.
- There is insufficient evidence comparing the harms of medical cannabis or cannabinoids to other pain management options, such as opioids.

## Background

Chronic pain is the primary cause of health care resource use and disability among working adults in North America and Western Europe^1 2^. The use of cannabis for the management of chronic pain is becoming increasingly common due to pressure to reduce opioid use, increased availability and changing legislation, shift in public attitudes and decreased stigma, and aggressive marketing^3 4^. The two most-studied cannabinoids in medical cannabis are delta-9-tetrahydrocannabinol (THC) and cannabidiol (CBD) ^5^. THC binds to cannabinoid receptors type 1 and 2, is an analog to the endogenous cannabinoid, anandamide, and has shown psychoactive, analgesic, anti-inflammatory, antioxidant, antipruritic, anti-spasmodic, and muscle-relaxant activities. CBD directly interacts with various ion channels to produce analgesic, anti-inflammatory, anti-convulsant and anxiolytic activities, without the psychoactive effect of THC^5^. Use of cannabis for therapeutic purposes, however, remains contentious due to its known and suspected harms^6-9^.

Though common adverse events caused by medical cannabis, including nausea, vomiting, headache, drowsiness, and dizziness, have been well documented in randomized controlled trials and reviews of randomized controlled trials, ^10 11^ less is known about potentially uncommon but serious adverse events, particularly events that may occur with longer durations of medical cannabis use, such as dependence, withdrawal symptoms, and psychosis^4 12-17^. Such adverse events are usually observed in large non-randomized studies that recruit larger numbers of patients and typically follow them for longer durations of time. Further, evidence from non-randomized studies may be more generalizable, since randomized controlled trials typically use strict eligibility criteria.

The objective of this systematic review and meta-analysis is to summarize the evidence on the risks and, when evidence on risk is not available, the prevalence of adverse events related to medical cannabis and cannabinoids from non-randomized studies. This evidence synthesis is part of the *BMJ Rapid Recommendations* project, a collaborative effort from the *MAGIC Evidence Ecosystem Foundation* (www.magicevidence.org) and the *BMJ*^18^. A guideline panel helped define the study question and selected adverse events for review. The adverse events of interest include psychiatric and cognitive adverse events, injuries and accidents, and dependence and withdrawal. It is one of four systematic reviews that together informed a parallel guideline on medical cannabis and cannabinoids published on bmj.com and the MAGICapp.^11 19-21^ A parallel systematic review addressed evidence from randomized trials.^11^

## Methods

We report our systematic review in accordance with the PRISMA Harms Checklist^22^. We registered the protocol for our review at OSF (https://osf.io/25bxf). ^22^

### Guideline panel involvement

A guideline panel helped define the study question and selected the adverse events for review. The panel included nine content experts (two general internists, two family physicians, a pediatrician, a physiatrist, a pediatric anesthesiologist, a clinical pharmacologist, and a rheumatologist), nine methodologists (five of whom are also front-line clinicians), and three people living with chronic pain (one of whom used cannabinoids for medical purposes).

### Patient and public involvement

Three patient partners were included as part of the guideline panel and contributed to the selection and prioritization of outcomes, protocol, and interpretation of review findings, and provided insight on values and preferences.

### Search

A medical librarian searched MEDLINE, EMBASE, PsychInfo, and Cochrane Central Register of Controlled Trials (CENTRAL) from inception to April 1, 2020, with no restrictions on language, for non-randomized studies reporting on harms or adverse events of medical cannabis or cannabinoids for chronic pain (Appendix 1). We scanned reference lists of relevant reviews to identify any eligible studies not retrieved by our electronic search and solicited content experts from our panel for unpublished studies.

### Study selection

Reviewers, working independently and in duplicate, reviewed titles and abstracts of search records and subsequently full texts of records found potentially eligible at the title and abstract screening stage. Reviewers resolved disagreements by discussion or by adjudication by a third reviewer (DZ).

We included all non-randomized studies that reported on any patient-important harm or adverse event associated with the use of any formulation of medical cannabis or cannabinoids in adults or children, living with chronic pain (pain lasting for ≥3 months) or a medical condition associated with chronic pain (i.e., fibromyalgia, arthritis, multiple sclerosis, neuropathy, inflammatory bowel disease, stroke, or advanced cancer) or that compared adverse events associated with medical cannabis or cannabinoids with another pharmacologic or non-pharmacologic intervention. Based on input from the guideline panel, we excluded studies in which patients used cannabis for less than 4 weeks because we anticipated that four weeks would be the minimum amount of time after which we would reasonably expect to observe potential serious or long term harms associated with medical cannabis. ^23^ We looked for explicit statements or evidence that patients were experiencing chronic pain. We excluded studies in which: (1) fewer than 25 patients used medical cannabis or cannabinoids, (2) patients did not suffer from chronic pain or a condition that commonly causes chronic pain or more than 20% of patients reported using medical cannabis or cannabinoids for a condition other than chronic pain, (3) patients were using medical cannabis for recreational reasons, (4) only surrogate measures of patient-important harms and adverse effects (e.g., performance on cognitive tests, lab values) were reported, and (5) systematic reviews and other types of studies that did not describe primary data.

### Data extraction and risk of bias

Reviewers, working independently and in duplicate and using a standardized and pilot-tested data collection form, extracted the following information from each eligible study: (1) study design, (2) patient characteristics (age, sex, condition/diagnosis), (3) characteristics of medical cannabis or cannabinoids (name of product, dose, and duration), and (4) number of patients that experienced adverse events, including all adverse events, serious adverse events, and withdrawal due to adverse events. We classified adverse events as serious based on the classification used in primary studies. For comparative studies, we collected results from models adjusted for confounders, when reported, and unadjusted models when results for adjusted models were not reported.

When studies reported the number of events rather than the number of patients experiencing adverse events, we only extracted the number of events if they were infrequent (the number of events accounted for less than 10% of the total number of study participants). For studies that reported on adverse events at multiple timepoints, we extracted data for the longest point of follow-up that included, at minimum, 80% of the patients recruited into the study. Reviewers resolved disagreements by discussion or by adjudication with a third reviewer (DZ).

We used the Cochrane-endorsed ROBINS-I tool to rate the risk of bias of studies as low, moderate, serious, or critical across seven domains: (1) bias due to confounding, (2) selection of patients into the study, (3) classification of the intervention, (4) bias due to deviations from the intended intervention, (5) missing data, (6) measurement of outcomes, and (7) selection of reported results. ^24^ Appendix 2 presents additional details on the assessment of risk of bias. Studies were rated at low risk of bias overall when all domains were at low risk of bias; moderate risk of bias if all domains were rated at low or moderate risk of bias; at serious risk of bias when all domains were rated either at low, moderate, or serious risk of bias; and at critical risk of bias when one or more domains were rated as critical.

### Data synthesis

In this review, we synthesize data on serious adverse events and adverse events that may emerge with longer duration of medical cannabis use for which data is typically not reported in randomized trials. Identified by a parallel BMJ Rapid Recommendations guideline panel as important, these patient-important outcomes included psychiatric and cognitive adverse events, injuries and accidents, and dependence and withdrawal. Data on all other adverse events reported in primary studies are available in an open-access database (https://osf.io/ut36z/).

Adverse events are reported as binary outcomes. For comparative studies, when possible, we present risk differences and associated 95% confidence intervals (95% CIs). Since there were only two eligible comparative studies each with different comparators, we did not perform meta-analysis. For single-arm studies, we pooled the proportion of patients experiencing adverse events of interest by first applying a Freeman-Tukey type arcsine square root transformation to stabilize the variance. Without this transformation, very high or very low prevalence estimates can produce confidence intervals that contain values lower than 0% or higher than 100%. All meta-analyses used DerSimonian-Laird random-effects models, which are conservative as they consider both within- and between-study variability. ^25-27^ We evaluated heterogeneity for all pooled estimates through visual inspection of forest plots and calculation of tau-squared (τ^2^), because some statistical tests of heterogeneity (I^2^ and Cochrane’s Q) can be misleading when sample sizes are large and CIs are therefore narrow. ^28^ For studies that reported estimates for all-cause adverse events and those deemed to be potentially related to cannabis use, we preferentially synthesized results for all adverse events.

For analyses for which we observed high clinical heterogeneity (i.e., substantial differences in the estimates of individual studies and minimal overlap in the confidence intervals), we presented results narratively.

We classified adverse events as serious based on the classification used in primary studies.

We performed tests of interaction to establish whether subgroups differed significantly from one another. We assessed the credibility of significant subgroup effects (test of interaction p⍰<⍰.05) using published criteria. ^29 30^

We performed all analyses using the ‘meta’ package in R (version 3.5.1, R Foundation for Statistical Computing). ^31^

### Certainty of evidence

We used the GRADE approach to rate the certainty of evidence^32 33^. Based on GRADE guidance for using the ROBINS-I tool, evidence starts at high certainty and is downgraded by one level when the majority of the evidence comes from studies at moderate risk of bias, two levels when the majority of the evidence comes from studies at high risk of bias, and three levels when the majority of the evidence comes from studies rated at critical risk of bias. ^32^ We additionally considered potential limitations due to indirectness if the population, intervention, or adverse events assessed in studies did not reflect the populations, interventions, or adverse events of interest, inconsistency if there was important unexplained differences in the results of studies, and imprecision if the upper and lower bounds of confidence intervals indicated appreciably different rates of adverse events. For assessing inconsistency and imprecision for the outcome all adverse events, based on feedback from the guideline panel, we deemed a 20% difference in the prevalence of all adverse evidence to be patient-important; a 10% difference for adverse events leading to discontinuation, serious adverse events, and psychiatric, cognitive, withdrawal and dependence, injuries; and a 3% difference for potentially fatal adverse events, such as suicides and motor vehicle accidents. We followed GRADE guidance for communicating our findings. ^34^ Guideline panel members interpreted the magnitude of adverse events and decided whether the observed prevalence of adverse events was sufficient to affect patients’ decisions to use medical cannabis or cannabinoids for chronic pain.

## Results

### Study selection

Our search yielded 17,178 unique records of which 39 were eligible for review (Figure 1, Appendix 3). ^35-73^ Appendix 4 presents studies excluded at the full-text screening stage and accompanying reasons for exclusion.

**Figure 1:**
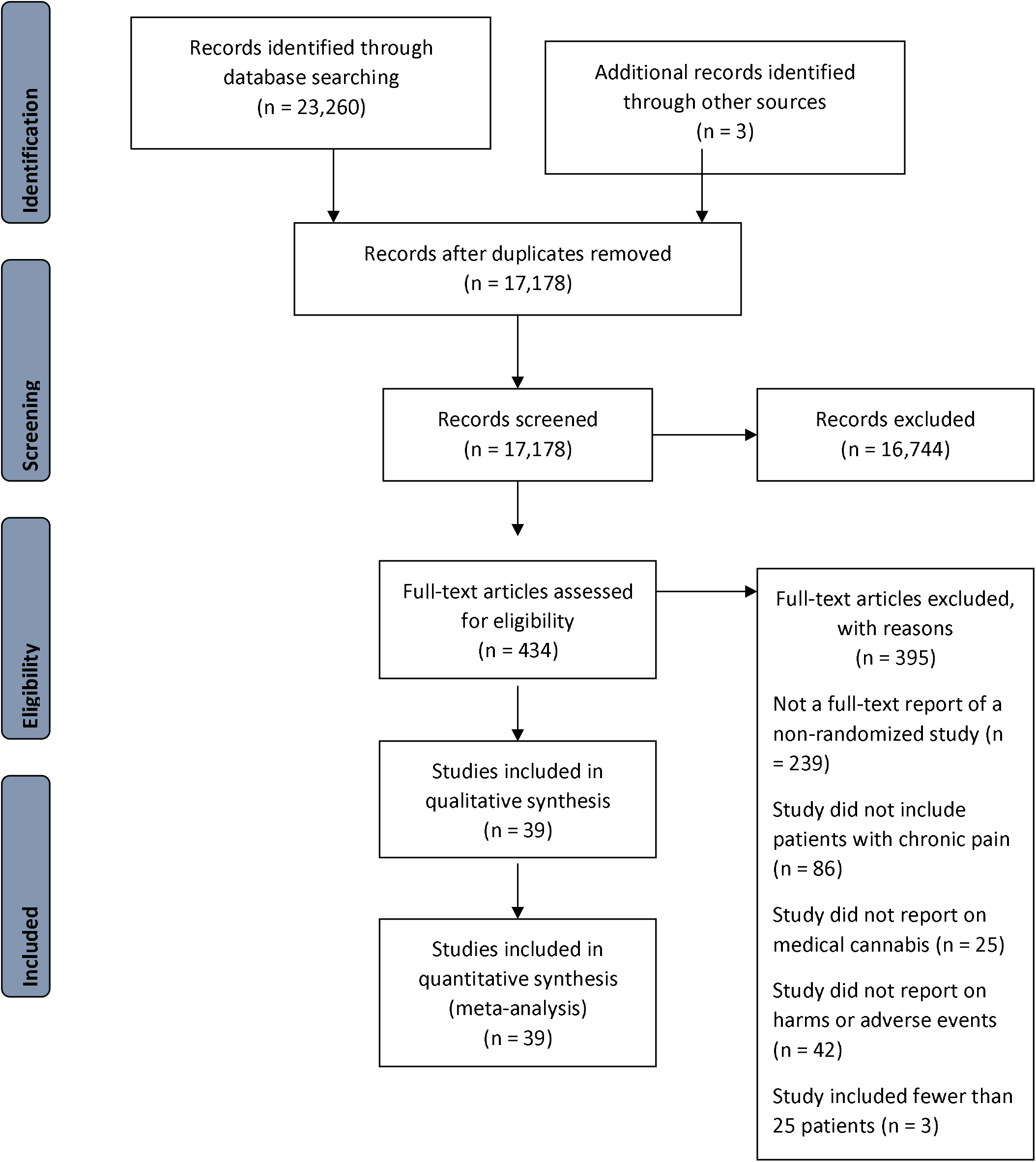
Study selection process.

### Description of studies

Studies included 12,143 adults living with chronic pain and included a median of 100 (IQR 34 to 361) participants (Table 1). Most studies (30/39; 76.9%) were longitudinal in design. Eighteen studies (46.2 %) were conducted in Western Europe, fourteen (35.9%) in North America, six (15.4%) in Israel, and two (5.1%) in the United Kingdom. Ten studies (25.6%) were funded by industry alone or industry in combination with government and institutional funds; the remainder were funded either by governments, institutions, or not-for-profit organizations (n=9; 23.1%), did not receive funds (n=3; 7.7%), or did not report funding information (n=17; 43.6%).

**Table 1:**
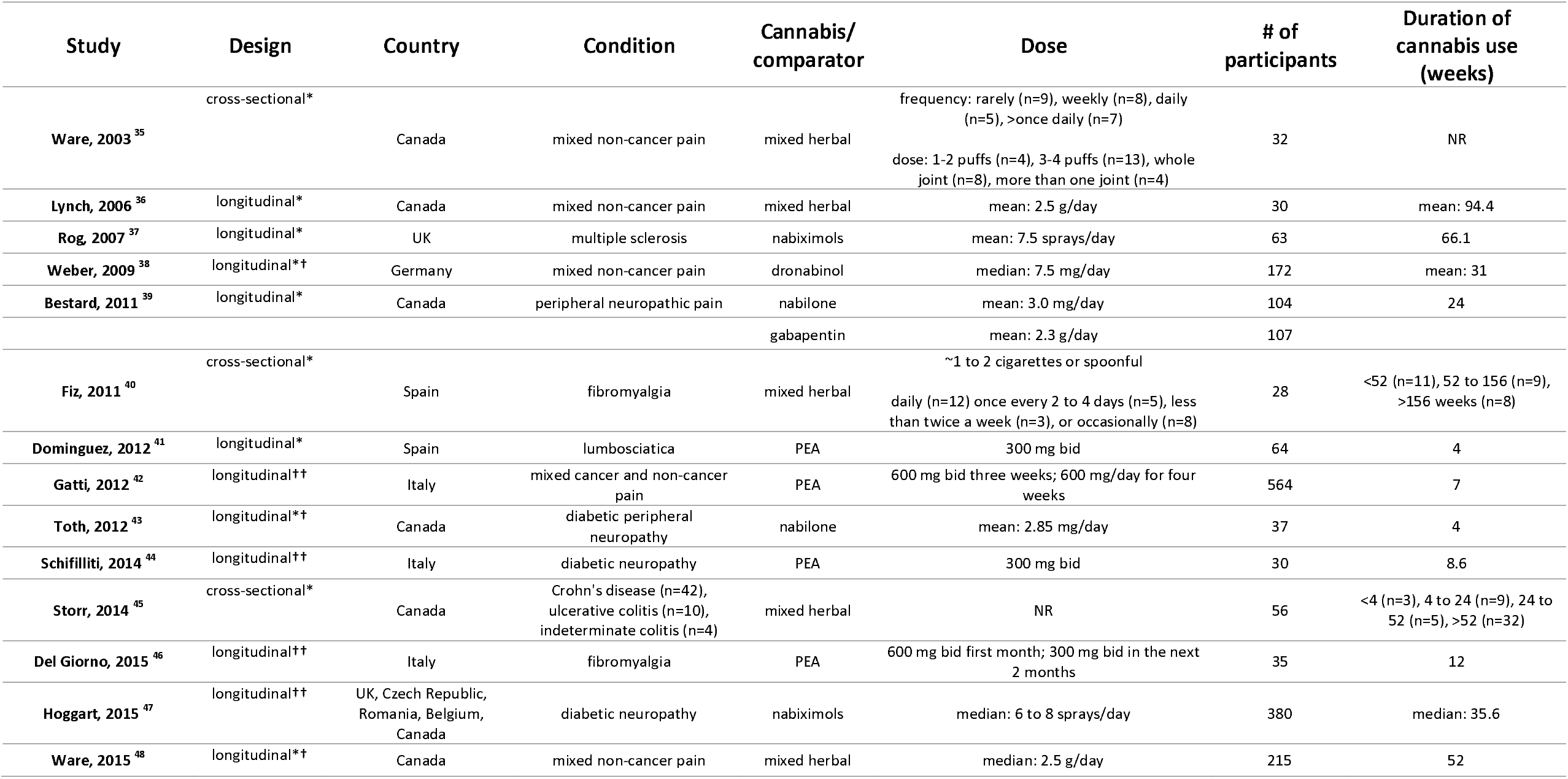
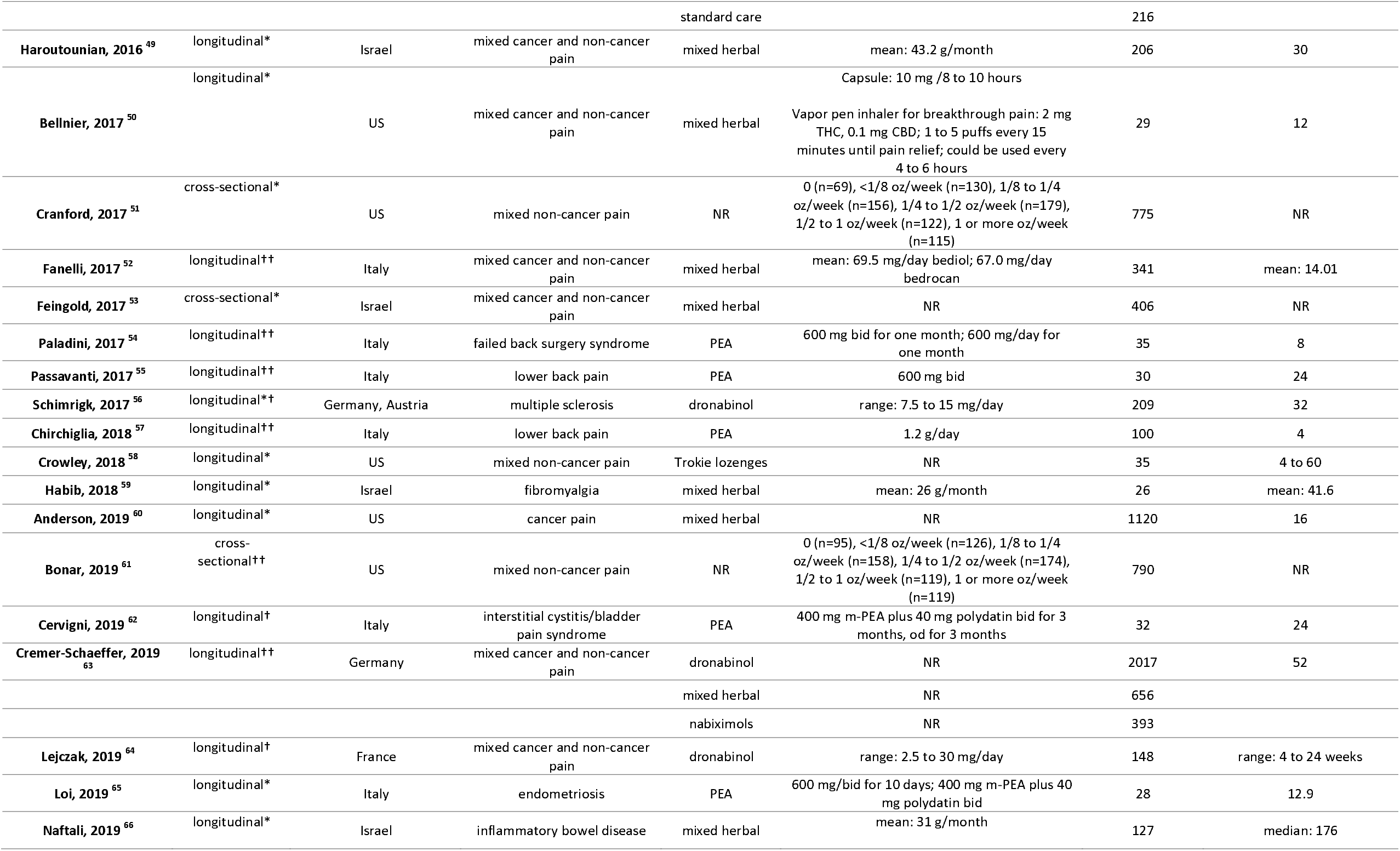
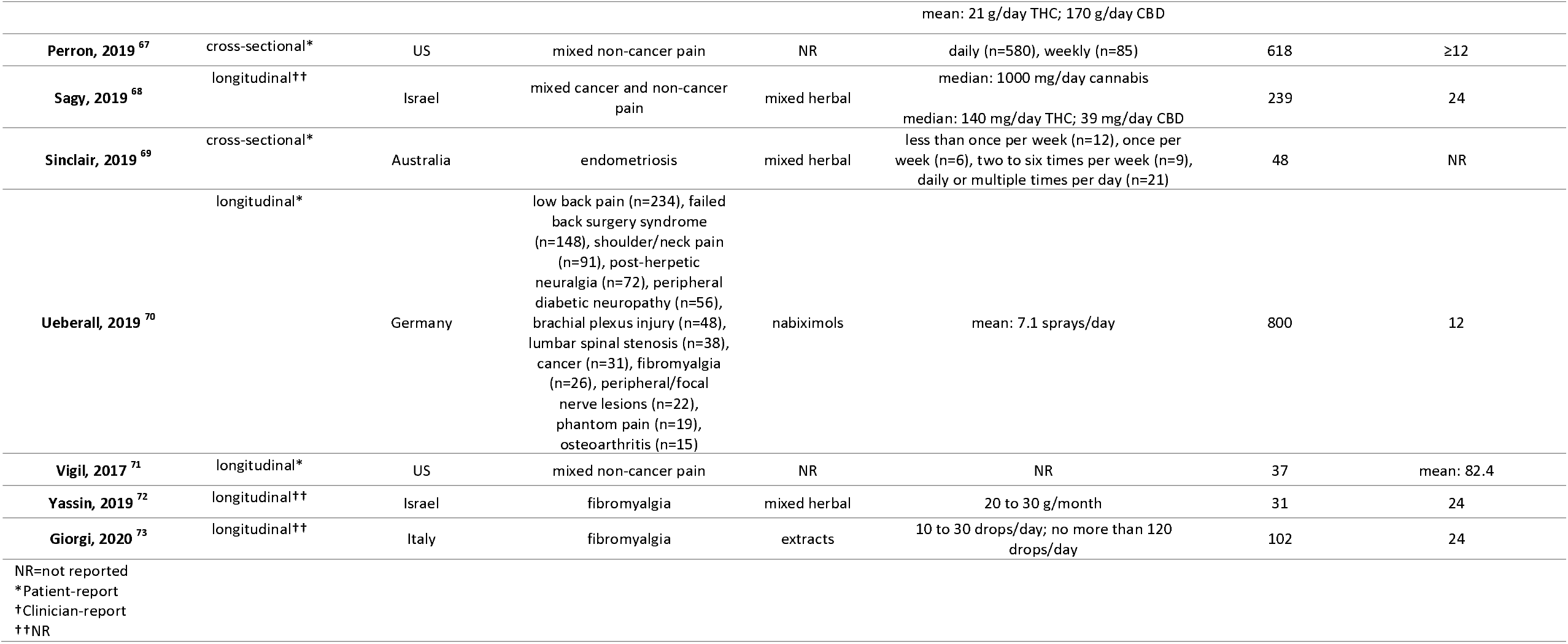
Study characteristics.

Thirty studies (76.9%) reported on people living with chronic non-cancer pain, eight (n=20.5%) with mixed cancer and non-cancer chronic pain, and one (2.6%) with chronic cancer pain. All studies reported on adults. Sixteen studies reported on mixed types of herbal cannabis (e.g., buds for smoking, vaporizing, and ingesting, hashish, oils, extracts, edibles), nine on palmitoylethanolamide (PEA), four each on nabiximols and dronabinol, two on nabilone, one each on Trokie lozenges and extracts, and four did not report the type of medical cannabis used. One study reported on three types of medical cannabis (dronabinol, nabiximols, and mixed herbal) separately. The median duration of medical cannabis use was 24 weeks (IQR 12.0 to 33.8 weeks). Two studies were comparative: one study compared nabilone with gabapentin and another compared herbal cannabis with standard care. ^39 48^ Studies reported a total of 525 unique adverse events.

### Risk of bias

Appendix 5 presents the risk of bias of included studies. We rated all results at critical risk of bias except for the comparative results from two studies, ^39 48^ which were rated at serious and moderate risk of bias. The primary limitation across studies was inadequate control for potential confounding either due to the absence of a control group or inadequate adjustment for confounders. A third of studies were rated at serious risk of bias for selection bias, primarily because they included prevalent users of medical cannabis. Such studies may underestimate the incidence of adverse events since patients that experience adverse events are more likely to discontinue medical cannabis early. Such studies may also include adverse events that may have been present at inception and that are unrelated to medical cannabis use.

### All adverse events

Twenty longitudinal and two cross-sectional studies, including 4,108 patients, reported the number of patients experiencing one or more adverse events. ^36-43 46 47 54 56-60 62 64 65 69 70 73^ Seven studies reported on PEA, five on mixed herbal cannabis, three each on nabilone and nabiximols, two on dronabinol, and one each on extracts and Trokie lozenges. The median duration of medical cannabis use was 24 weeks [IQR 12 to 32]. We observed substantial unexplained heterogeneity and so summarize the results descriptively (Appendices 6 to 9). The prevalence of any adverse event ranged between 0% to 92.1%. Studies with less than 24 weeks of cannabis use typically reported fewer adverse events than those with more than 24 weeks. Patients using PEA experienced no adverse events. The evidence was overall very uncertain due to risk of bias and inconsistency.

One study suggested that nabilone may reduce the risk of adverse events compared to gabapentin (−13.1%; 95% CI -26.2 to 0), but the certainty of evidence was very low due to risk of bias and imprecision (Table 2).

**Table 2:** Prevalence of adverse events from non-comparative studies.

| Outcome | Number of studies | Number of participants | Duration of follow-up (weeks) | Prevalence % (95% CI) | I<sup>2</sup> (%) | Certainty | Reasons for downgrading |
| --- | --- | --- | --- | --- | --- | --- | --- |
| **All adverse events** | 22 | 4,108 | 4 to 94 | The prevalence of adverse events ranged between 0% to 92.1%. Studies with less than 24 weeks of cannabis use typically reported fewer adverse events than those with more than 24 weeks. Patients using PEA experienced no adverse events. The evidence was overall very uncertain due to risk of bias and inconsistency. |  | very low | risk of bias (3 levels), inconsistency |
| **Adverse events causing discontinuation** | 20 | 6,509 | 4 to 66 | The prevalence of discontinuations due to adverse events ranged between 0% to 27.0%. Studies with less than 24 weeks of cannabis use typically reported fewer discontinuations than those with more than 24 weeks. Patients using PEA experienced no adverse events. The evidence was overall very uncertain due to risk of bias and inconsistency. |  | very low | risk of bias (3 levels), inconsistency |
| **Serious adverse events** | 24 | 4,273 | 4 to 94 | 1.2 (0.1 to 3.1) | 91 | very low | risk of bias (3 levels) |
| **Psychiatric adverse events** |  |  |  |  |  |  |  |
| Psychiatric disorder | 4 | 1,458 | 12 to 66 | 13.5 (2.6 to 30.6) | 98 | very low | risk of bias (3 levels), inconsistency, imprecision |
| Suicide | 1 | 215 | 52 | 0 (0 to 0.8) | NA | very low | risk of bias (3 levels) |
| Suicidal thoughts | 1 | 3,066 | 52 | 0.1 (0 to 0.5) | 44 | very low | risk of bias (3 levels) |
| Depression | 6 | 4,144 | 12 to 66 | 1.7 (0.9 to 2.7) | 71 | very low | risk of bias (3 levels) |
| Mania | 1 | 215 | 52 | 0.5 (0 to 2) | NA | very low | risk of bias (3 levels) |
| Hallucinations | 6 | 3,583 | 24 to 66 | 0.5 (0.1 to 1.3) | 69 | very low | risk of bias (3 levels) |
| Delusions | 4 | 3,281 | 52 | 0.4 (0.2 to 0.6) | 0 | very low | risk of bias (3 levels) |
| Paranoia | 3 | 277 | 52 to 94; one cross-sectional study | 5.6 (0 to 19.2) | 85 | very low | risk of bias (3 levels), inconsistency, imprecision |
| Anxiety | 5 | 1,695 | 12 to 94;<br>two cross-sectional studies | 7.4 (0 to 26.9) | 99 | very low | risk of bias (3 levels), imprecision |
| Euphoria | 7 | 4,501 | 4 to 66 | 2.1 (0.9 to 3.8) | 96 | very low | risk of bias (3 levels) |
| **Cognitive adverse events** |  |  |  |  |  |  |  |
| Memory impairment | 6 | 4,484 | 4 to 176 | 5.3 (2.1 to 9.6) | 96 | very low | risk of bias (3 levels) |
| Confusion | 7 | 1,654 | 4 to 176 | 1.8 (0.3 to 4.2) | 81 | very low | risk of bias (3 levels) |
| Disorientation | 6 | 4,485 | 12 to 52 | 1.6 (0.6 to 3.0) | 88 | very low | risk of bias (3 levels) |
| Attention disorder or deficit | 8 | 5,477 | 12 to 82 | 3.4 (1.3 to 6.3) | 95 | very low | risk of bias (3 levels) |
| **Accidents and injuries** |  |  |  |  |  |  |  |
| Falls | 1 | 215 | 52 | 2.3 (0.7 to 4.9) | NA | very low | risk of bias (3 levels) |
| Motor vehicle accidents | 1 | 215 | 52 | 0.5 (0 to 2.0) | NA | very low | risk of bias (3 levels) |
| **Dependence and withdrawal** |  |  |  |  |  |  |  |
| Dependence | 3 | 1,824 | 12; one cross-sectional study | 4.4 (0.0 to 19.9) | 99 | very low | risk of bias (3 levels), inconsistency, imprecision, indirectness |
| Withdrawal syndrome | 2 | 424 | 32 to 52 | 2.1 (0 to 8.2) | 89 | very low | risk of bias (3 levels), indirectness |
| Withdrawal symptoms | 1 | 618 | NA; cross-sectional | 67.8 (64.1 to 71.4) | NA | very low | risk of bias (3 levels), indirectness |

**Table 3:**
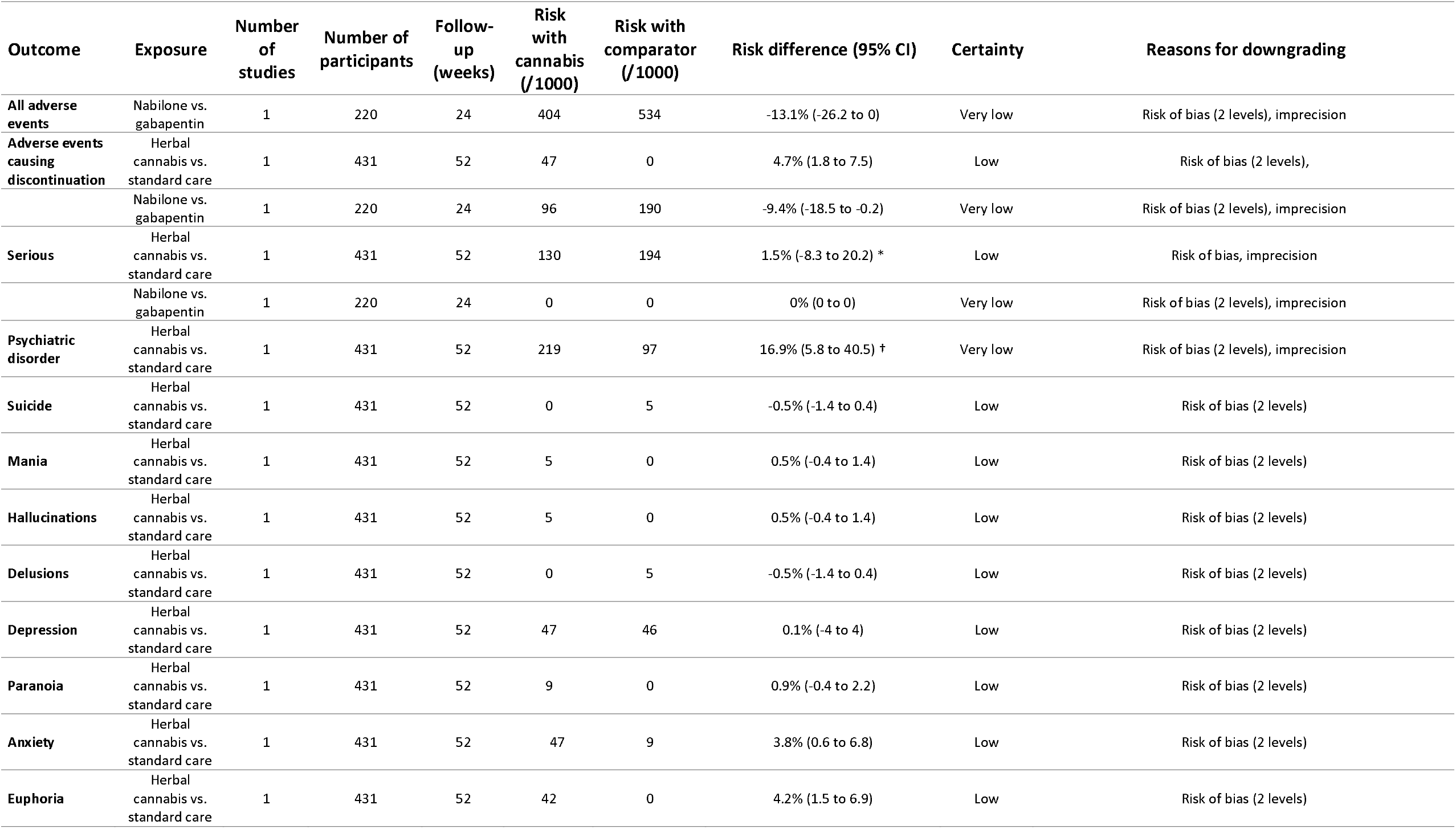
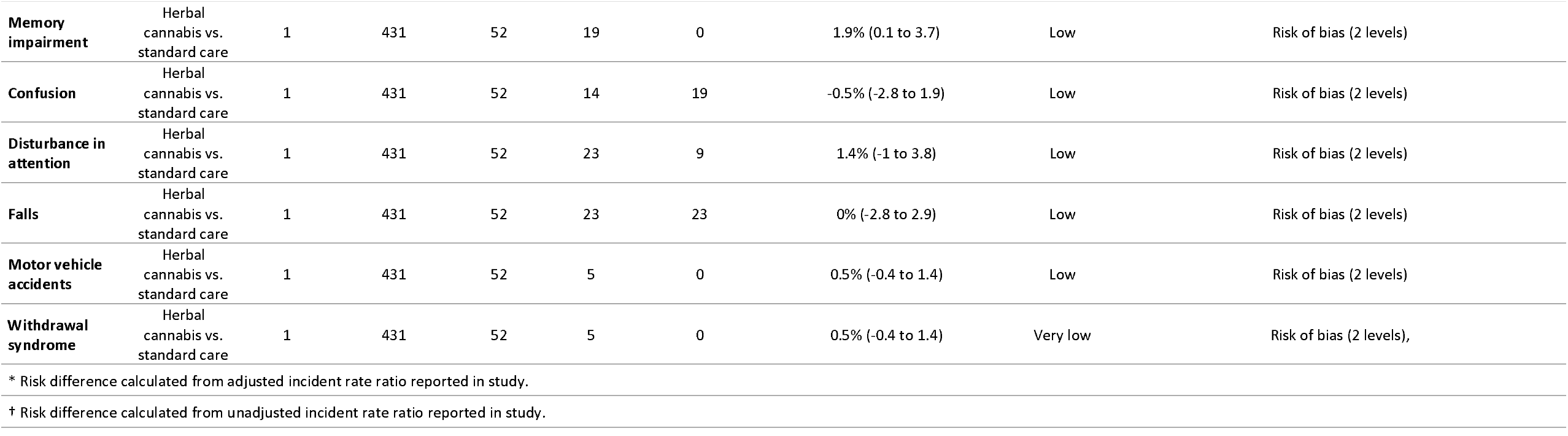
Risk differences for adverse events from comparative studies.

### Adverse events leading to discontinuation

Twenty longitudinal studies, including 6,509 patients, reported on the number of patients that discontinued medical cannabis or cannabinoids due to adverse events. ^37 39 41-44 46-49 52 54 56 57 59 62 63 65 70 73^ Eight studies reported on PEA, four studies on mixed herbal cannabis, three on nabiximols, two on nabilone, and one each on dronabinol and extracts, and one study did not report the type of medical cannabis used by patients. The median duration of cannabis use was 24 weeks [IQR 8.6 to 32]. We observed substantial unexplained heterogeneity and so summarize the results descriptively (Appendices 10 to 12). The prevalence of discontinuations due to adverse events ranged between 0% to 27.0%. Studies with less than 24 weeks of cannabis use typically reported fewer discontinuations than those with more than 24 weeks. Patients using PEA experienced no adverse events. The evidence was overall very uncertain due to risk of bias and inconsistency.

One study suggested herbal cannabis may increase the risk of adverse events leading to discontinuation compared to standard care without cannabis (4.7%; 95% CI 1.8 to 7.5). Another study suggested that nabilone may reduce the risk of adverse events leading to discontinuation compared to gabapentin (−9.4%; 95% CI -18.5 to -0.2). The certainty of evidence was low to very low due to risk of bias and imprecision.

### Serious adverse events

Twenty-two longitudinal and two cross-sectional studies, including 4,273 patients, reported on the number of patients experiencing one or more serious adverse events. ^35-37 39-43 46 48 49 52 54-60 62 65 70 71 73^ Eight studies reported on mixed herbal cannabis, eight on PEA, two each on nabilone and nabiximols each, and one study each on dronabilon, extracts, and Trokie lozenges, and one study did not report the type of cannabis used. The median duration of medical cannabis or cannabinoid use was 24 weeks (IQR 12 to 32), and few patients experienced serious adverse events (1.2%; 95% CI 0.1 to 3.1; I^2^ =91%) (Figure 2) (Appendices 13 to 15). There was a statistically significant subgroup effect across different types of medical cannabis though serious adverse events appeared consistently uncommon among different types (low credibility). The certainty of evidence was very low overall due to serious risk of bias.

**Figure 2:**
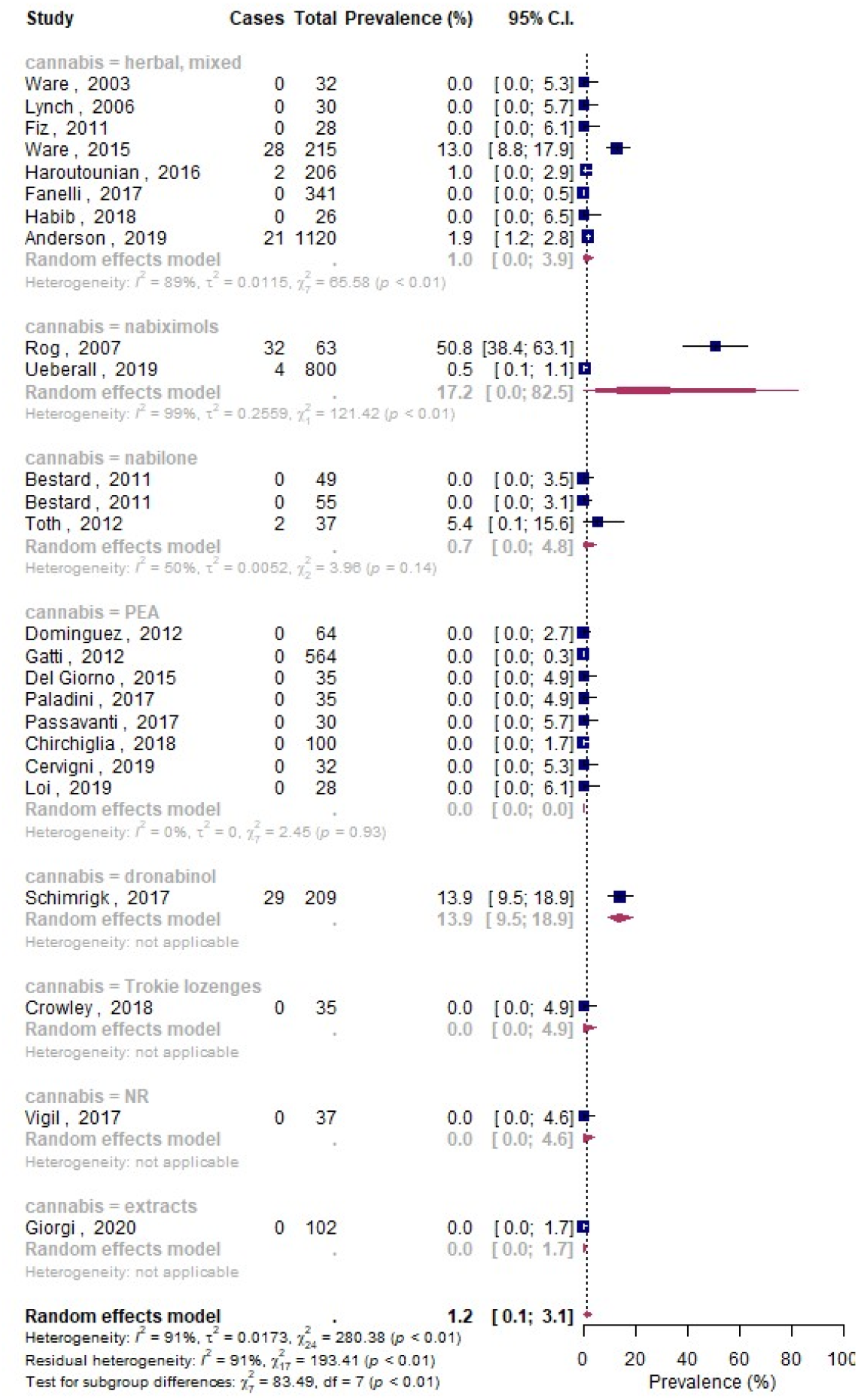
Forest plot of the meta-analysis for all adverse events stratified by type of medical cannabis.

One study suggested herbal cannabis increased the risk of serious adverse events compared to standard care without cannabis (1.5%; 95% CI -8.3 to 20.2). Another study found use of nabilone vs. gabapentin showed no difference in the risk of serious adverse events. The certainty of evidence was low to very low for both studies due to risk of bias and imprecision.

### Psychiatric adverse events

Eleven longitudinal and two cross-sectional studies, including 6,600 patients, reported on any psychiatric adverse events, including psychiatric disorders, suicide, suicidal thoughts, depression, mania, hallucinations, delusions, paranoia, anxiety, and euphoria (Appendices 16 to 25). ^35-37 43 47 48 60 63 67 68 70^ Five studies reported on mixed herbal cannabis, four on nabiximols, one each on dronabinol, nabilone, and mixed types and one study did not specify the type of medical cannabis. The median duration of cannabis use across studies was 52 weeks (IQR 20 to 52). Approximately one in seven medical cannabis users experienced one or more psychiatric disorders or adverse events (13.5%; 95% CI 2.6 to 30.6; I^2^ =98%). The most frequently occurring psychiatric adverse events were paranoia (5.6%; 9% CI 0 to 19.2; I^2^ =85%) and anxiety (7.4%; 95% CI 0 to 26.9; I^2^ =99%). The certainty of evidence was very low due to risk of bias, inconsistency (for psychiatric disorders and paranoia), and imprecision (for psychiatric disorder, paranoia, and anxiety).

One study suggested that herbal cannabis may result in a trivial to moderate increase in the risk for psychiatric disorders, mania, hallucinations, depression, paranoia, anxiety, and euphoria and a reduction in the risk for suicides and delusions, compared with standard care without cannabis, though the certainty of evidence was low to very low due to risk of bias and imprecision.

### Cognitive and attentional adverse events

Eleven longitudinal studies, including 6,257 patients, reported on cognitive adverse events, including memory impairment, confusion, disorientation, and impaired attention (Appendices 26 to 29). ^35-37 43 47 48 60 63 67 68 70^ Five studies reported on herbal cannabis, three on nabiximols, three on mixed types of cannabis, and one each on dronabinol and nabilone. The median duration of cannabis use was 52 weeks (IQR 24 to 52). The prevalence of cognitive adverse events ranged from 1.6% (95% CI 0.6 to 3.0; I^2^ =88%) to 5.3% (95% CI 2.1 to 9.6; I^2^ =96%) for disorientation and memory impairment, respectively. The certainty of evidence was very low due to risk of bias.

One study suggests herbal cannabis may slightly increase the risk for memory impairment and disturbances in attention compared to standard care without cannabis, but reduce the risk for confusion, though the certainty of evidence was low to very low due to risk of bias and imprecision.

### Accidents and injuries

One longitudinal study, including 431 patients, reported on accidents and injuries in patients using mixed herbal cannabis for 52 weeks (Appendices 30 & 31). ^48^ This study suggests herbal cannabis used for medical purposes may slightly increase the risk of motor vehicle accidents (0.5%; 95% CI -0.4 to 1.4) but may not increase the risk of falls (0%; 95% CI -2.8 to 2.9). The certainty of evidence was low due to risk of bias.

### Dependence and withdrawal

Four longitudinal and one cross-sectional study, including 2,248 patients, reported on dependence-related adverse events, including dependence (one study reported on ‘abuse’ based on unspecified criteria, one study reported on ‘problematic use’ using the Alcohol Use Disorder and Associated Disabilities Interview Schedule–Diagnostic and Statistical Manual of Mental Disorders–Fourth Edition (AUDADIS-IV) ^74^, and one study reported on ‘dependence’ using the Alcohol, Smoking, and Substance Involvement Screening Test^75^), withdrawal symptoms (defined as one or moderate or severe withdrawal symptoms including sleep difficulties, anxiety, irritability, and appetite disturbance), and withdrawal syndrome (two studies that used unspecified criteria) (Appendices 32 to 34). ^48 53 56 67 70^ Two studies reported on herbal cannabis, one each on nabiximols and nabilone, and one did not specify type of medical cannabis used by patients. Follow-up ranged from 12 to 52 weeks. Though dependence and withdrawal syndrome were uncommon with a prevalence of 4.4% (95% CI 0.0 to 19.9; I^2^=99%) and 2.1% (95% CI 0 to 8.2; I^2^ =89%), respectively, withdrawal symptoms were common (67.8%; 95% CI 64.1 to 71.4). The certainty of evidence was very low due to risk of bias, inconsistency, imprecision (for dependence), and indirectness due to definitions of outcomes in studies were too vague to confidently distinguish between dependence, addiction, withdrawal symptoms, and withdrawal syndrome.

One study suggested that herbal cannabis compared to standard care may slightly increase the risk of withdrawal syndrome (0.5%; 95% CI -0.4 to 1.4) but the certainty of evidence was low due to risk of bias.

## Discussion

### Main findings

Our systematic review and meta-analysis provides evidence that adverse events are common among people living with chronic pain who use medical cannabis or cannabinoids, with approximately one in four experiencing at least one adverse event—though the certainty of evidence is very low and the true prevalence of adverse events may be substantially different. In contrast, serious adverse events, adverse events leading to discontinuation, cognitive adverse events, accidents and injuries, and dependence and withdrawal syndrome are uncommon. We compared studies with <24 weeks and ≥ 24 weeks cannabis use and found more adverse events reported among studies with longer follow-up. This may be explained by increased tolerance (tachyphylaxis) with prolonged exposure, necessitating increases in dosage with consequent increased risk of harms. PEA, compared to other formulations of medical cannabis, may result in the fewest adverse events. Though adverse events appear to be common, few patients discontinued medical cannabis due to adverse events suggesting that most adverse events are transient and/or outweighed by perceived benefits.

Our review represents the most comprehensive review of evidence from non-randomized studies addressing adverse events of medical cannabis or cannabinoid use in people living with chronic pain. While several previous reviews have summarized the evidence on short-term and common adverse events of medical cannabis reported in randomized trials, such as oral discomfort, dizziness, and headaches, our review focuses on serious and rare adverse events—the choice of which was informed by a panel including patients, clinicians, and methodologists—and non-randomized studies, which can follow larger numbers of patients for longer periods of time and thus may detect adverse events that are infrequent or that are associated with longer durations of cannabis use. ^10 76-80^ A parallel systematic review of evidence from randomized controlled trials found no evidence to inform long-term harms of medical cannabis as no eligible trial followed patients for more than 5.5 months. ^11^ One previously published review that included non-randomized studies searched the literature until 2007, included studies exploring medical cannabis for any indication (excluding synthetic cannabinoids) of which only two enrolled people living with chronic pain. ^12^ The review also did not synthesize adverse event data from non-randomized studies. ^12^ Unlike previous reviews, we focused exclusively on medical cannabis for chronic pain and excluded recreational cannabis, because cannabis used for recreational purposes often contains higher concentrations of tetrahydrocannabinol (THC) than medical cannabis. We also focused on chronic pain because this patient population may be susceptible to different adverse events. Depression and anxiety, for example, are commonly occurring comorbidities of chronic pain, which may be exacerbated by cannabis. ^15-17^

### Strengths and limitations

Strengths of this systematic review and meta-analysis include a comprehensive search for non-randomized studies, explicit eligibility criteria, screening of studies and collection of data in duplicate to increase reliability, and use of the GRADE approach to evaluate the certainty of evidence.

Our review is limited by the non-comparative design of most studies, which precludes confident inferences regarding the proportion of adverse events that can be attributed to medical cannabis or cannabinoids and the magnitude by which medical cannabis may increase or decrease the risk of adverse events compared to other pain management options. Though adverse events appear common among medical cannabis users, it is possible that other management options for chronic pain, particularly opioids, may be associated with more (and more severe) adverse events. ^81^Partly due to the non-comparative design of most studies, nearly all results included in our review were at serious or critical risk of bias for confounding, either due to the absence of a control group or due to insufficient adjustment for important confounders. Further, a third of studies were at high risk of selection bias, primarily because they included prevalent cannabis users. In such studies, the prevalence of adverse events may be underestimated. Our review provides limited evidence on the harms of medical cannabis beyond one year of use since most studies reported adverse events for less than one year of follow-up.

We observed some inconsistency for many adverse events of interest and substantial inconsistency for all adverse events and adverse events leading to discontinuation. We downgraded the certainty of evidence when we observed important inconsistency and we did not present estimates from meta-analyses for all adverse events and adverse events leading to discontinuation due to substantial inconsistency.

Sixteen of 39 studies reported on herbal medical cannabis, some of which were consumed by smoking or vaporizing, and may be associated with different adverse events (e.g. respiratory) than other formulations of medical cannabis. We attempted to perform subgroup analyses based on the type of medical cannabis. Results for subgroups, however, lacked credibility due to inconsistency and/or imprecision.

Clinicians and patients may be more inclined to use medical cannabis or cannabinoids for pain relief if adverse events are mild; however, the evidence on whether adverse events are transient, life threatening, or the extent to which they impact quality of life is limited. While more than half of studies reported on the proportion of adverse events that were serious, criteria for ascertaining severity were rarely reported. None of the included studies reported the duration for which patients experienced adverse events. Further, most primary studies did not report adequate details on methods for the ascertainment of adverse events, including definitions or diagnostic criteria. The two studies that reported on withdrawal syndrome, for example, did not provide diagnostic criteria. ^48 56^ However, the DSM-5 requires ≥3 of 7 withdrawal symptoms to be present within a week of stopping cannabis use to meet a diagnosis of cannabis withdrawal syndrome. ^82^ It is therefore reasonable that people living with chronic pain that use medical cannabis would be more likely to experience withdrawal symptoms vs. withdrawal syndrome.

While children and youth account for approximately 15% of all chronic pain patients, we did not identify any evidence addressing the harms of medical cannabis in this population. ^83^ As such, the extent to which our findings are generalizable to pediatric populations is uncertain. Although there is evidence that cannabis use during youth is associated with increased risk of acute psychotic disorders, particularly acute psychosis, ^84^ such studies have explored use of recreational cannabis that contains greater amounts of THC than is typically seen in medical preparations. Further, the population of patients with chronic pain on which the studies report may not be representative of all patients with chronic pain— particularly rare conditions that cause chronic pain.

Finally, we excluded studies from meta-analyses when they did not explicitly report the adverse events of interest to our panel members. This may have overestimated the prevalence of adverse events if the adverse events of interest were not observed in the studies in which they were not reported. This was, however, not possible to confirm because methods for the collection and reporting of adverse event data across studies were variable (e.g., active monitoring vs. passive surveillance; collecting data on specific adverse events vs. all adverse events) and poorly described in study reports.

### Implications

Our systematic review and meta-analysis shows that evidence regarding long-term and serious harms of medical cannabis or cannabinoids is insufficient—an issue with important implications for patients and clinicians considering this management option for chronic pain. While the evidence suggests that adverse events are common in patients using medical cannabis for chronic pain, serious adverse events appear uncommon, which suggests that the potential benefits of medical cannabis or cannabinoids (although very modest) may outweigh potential harms for some patients. ^11 21^

Clinicians and patients considering medical cannabis should be aware that more adverse events were reported among studies with longer follow-up, necessitating long term follow-up of patients and re-evaluation of pain treatment options. Our findings also have implications for the choice of medical cannabis. We found PEA, for example, to consistently be associated with few or no adverse events across studies, though the evidence on the efficacy of PEA is limited. ^11^

We found very limited evidence comparing medical cannabis or cannabinoids with other pain management options. Other pharmacological treatments for chronic pain, such as gabapentinoids, antidepressants, and opioids, may be associated with more (and more serious) adverse events. ^85-87^ To guide patients’ and clinicians’ decisions on medical cannabis for chronic pain, future research should compare the harms of medical cannabis and cannabinoids with other pain management options, including opioids, ideally beyond one year of use, and adjust results for confounders. Future research could also explore whether the harms of medical cannabis vary depending on the type of chronic pain.

Our review highlights the need for standardization of reporting of adverse events in non-randomized studies since such studies represent a critical source of data on long-term and infrequently occurring harms. To enhance the interpretability of adverse event data, future studies should also report the duration and severity of adverse events, since these factors are important to patients’ decisions.

A valuable output of our systematic review is an open-source database of over 500 unique adverse events reported to date in non-randomized studies of medical cannabis or cannabinoids for chronic pain with corresponding assessments of risk of bias. This database was compiled in duplicate by trained and calibrated data extractors and is freely available to those interested in further analyzing the prevalence of different types of adverse or to those interested in expanding the database to include adverse events in patients using medical cannabis or cannabinoids for other indications.

## Conclusion

Our systematic review and meta-analysis found very low certainty evidence that suggests that adverse events are common among people living with chronic pain using medical cannabis or cannabinoids, but that serious adverse events, adverse events causing discontinuation, cognitive adverse events, motor vehicle accidents, falls, and dependence and withdrawal syndrome are uncommon. We also found very low certainty evidence that longer duration of use was associated more adverse events and that PEA, compared with other types of medical cannabis, may result in few or no adverse events. Future research should compare the risks of adverse events of medical cannabis and cannabinoids with alternative pain management options, including opioids, and adjust for potential confounders.

## Data Availability

Data are available upon reasonable request to the principal author

https://osf.io/ut36z/

## Abbreviations

CENTRAL: Cochrane Central Register of Controlled Trials
PEA: Palmitoylethanolamide
THC: tetrahydrocannabinol

## Acknowledgements

We thank the members of the Rapid Recommendations panel for critical feedback on the selection of the adverse events of interest. We thank James MacKillop, PhD, for his help with the interpretation of problematic cannabis use, abuse, dependance and withdrawal syndrome within studies.

