## Supplemental File for "Long-term and serious harms of medical cannabis and cannabinoids for chronic pain: A systematic review of non-randomized studies"

**Appendix**

Dr. Jason Busse

### Appendix 1: Search strategy

| MEDLINE | 10649 |
| --- | --- |
| EMBASE | 6382 |
| Central | 2426 |
| PsycInfo | 3801 |
| Subtotal | 23260 |
| -dupes | -6085 |
| Total | 17175 |

April 1, 2020

Database: OVID Medline Epub Ahead of Print, In-Process & Other Non-Indexed Citations, Ovid MEDLINE(R) Daily and Ovid MEDLINE(R) 1946 to Present

Search Strategy:

--------------------------------------------------------------------------------

1	Epidemiologic Studies/ (8256)

2	exp Case-Control Studies/ (1067341)

3	exp Cohort Studies/ (1974212)

4	Case control.tw. (123081)

5	(cohort adj (study or studies)).tw. (199133)

6	Cohort analy$.tw. (7799)

7	(Follow up adj (study or studies)).tw. (48708)

8	(observational adj (study or studies)).tw. (103255)

9	Longitudinal.tw. (239715)

10	Retrospective.tw. (515751)

11	Cross sectional.tw. (342224)

12	Cross-sectional studies/ (322752)

13	or/1-12 (2953281)

14	exp animals/ not humans.sh. (4685189)

15	13 not 14 (2889789)

Annotation: SIGN observational studies filter

16	randomized controlled trial.pt. (503041)

17	controlled clinical trial.pt. (93591)

18	randomized.ab. (474985)

19	placebo.ab. (206552)

20	drug therapy.fs. (2191450)

21	randomly.ab. (330409)

22	trial.ab. (500400)

23	groups.ab. (2028909)

24	or/16-23 (4670111)

25	exp animals/ not humans.sh. (4685189)

26	24 not 25 (4048339)

Annotation: Cochrane HSSS RCT filter

27	15 or 26 (6033576)

Annotation: study design filter broad

28	Cannabis/ (8968)

29	exp cannabinoids/ or cannabidiol/ or cannabinol/ or dronabinol/ (13810)

30	Endocannabinoids/ (5630)

31	exp Receptors, Cannabinoid/ (9240)

32	(Cannabis or cannabinol or cannabinoid* or cannabidiol or bhang or cannador or charas or ganja or ganjah or hashish or hemp or marihuana or marijuana or nabilone or cesamet or cesametic or ajulemic acid or cannabichromene or cannabielsoin or cannabigerol or tetrahydrocannabinol or dronabinol or levonantradol or nabiximols or palmidrol or tetrahydrocannabinolic acid or tetrahydro cannabinol or marinol or tetranabinex or sativex or endocannabinoid*).mp. (54925)

33	or/28-32 (54925)

Annotation: strategy from 2020 cannabis review

34	27 and 33 (16307)

Annotation: cannabis AND study design filter

35	exp "Drug-Related Side Effects and Adverse Reactions"/ (114376)

36	(ae or to or po or co).fs. (3890270)

37	(safe or safety).ti,ab. (758301)

38	side effect$.ti,ab. (243706)

39	((adverse or undesirable or harms$ or serious or toxic) adj3 (effect$ or reaction$ or event$ or outcome$)).ti,ab. (501888)

40	exp Product Surveillance, Postmarketing/ (15237)

41	adverse drug reaction reporting systems/ (7463)

42	clinical trials, phase iv/ (295)

43	exp Poisoning/ (156177)

44	exp Substance-Related Disorders/ (274845)

45	Abnormalities, Drug-Induced/ (14514)

46	Drug Monitoring/ (20599)

47	exp Drug Hypersensitivity/ (45642)

48	(toxicity or complication$ or noxious or tolerability).ti,ab. (1298802)

49	or/35-48 (5596308)

Annotation: OVID AE filter

50	34 and 49 (10649)

Annotation: Study design filter AND Cannabis AND AE Filter (broad)

Database: Embase <1974 to 2020 March 31>

Search Strategy:

--------------------------------------------------------------------------------

1	cannabis/ (33859)

2	exp cannabinoid/ (65694)

3	medical cannabis/ (2104)

4	exp cannabinoid receptor/ (14557)

5	exp endocannabinoid/ (8589)

6	(Cannabis or cannabinol or cannabinoid* or cannabidiol or bhang or cannador or charas or ganja or ganjah or hashish or hemp or marihuana or marijuana or nabilone or cesamet or cesametic or ajulemic acid or cannabichromene or cannabielsoin or cannabigerol or tetrahydrocannabinol or dronabinol or levonantradol or nabiximols or palmidrol or tetrahydrocannabinolic acid or tetrahydro cannabinol or marinol or tetranabinex or sativex or endocannabinoid*).mp. [mp=title, abstract, heading word, drug trade name, original title, device manufacturer, drug manufacturer, device trade name, keyword, floating subheading word, candidate term word] (86550)

7	or/1-6 (87843)

Annotation: cannabis

8	clinical study/ (154879)

9	case control study/ (153658)

10	family study/ (26012)

11	longitudinal study/ (137463)

12	retrospective study/ (897628)

13	prospective study/ (590879)

14	randomized controlled trials/ (176633)

15	13 not 14 (584662)

16	cohort analysis/ (564001)

17	(Cohort adj (study or studies)).mp. [mp=title, abstract, heading word, drug trade name, original title, device manufacturer, drug manufacturer, device trade name, keyword, floating subheading word, candidate term word] (296961)

18	(Case control adj (study or studies)).mp. [mp=title, abstract, heading word, drug trade name, original title, device manufacturer, drug manufacturer, device trade name, keyword, floating subheading word, candidate term word] (211490)

19	(follow up adj (study or studies)).mp. [mp=title, abstract, heading word, drug trade name, original title, device manufacturer, drug manufacturer, device trade name, keyword, floating subheading word, candidate term word] (65948)

20	(observational adj (study or studies)).mp. [mp=title, abstract, heading word, drug trade name, original title, device manufacturer, drug manufacturer, device trade name, keyword, floating subheading word, candidate term word] (242526)

21	(epidemiologic$ adj (study or studies)).mp. [mp=title, abstract, heading word, drug trade name, original title, device manufacturer, drug manufacturer, device trade name, keyword, floating subheading word, candidate term word] (109669)

22	(cross sectional adj (study or studies)).mp. [mp=title, abstract, heading word, drug trade name, original title, device manufacturer, drug manufacturer, device trade name, keyword, floating subheading word, candidate term word] (385983)

23	or/8-12,15-22 (2808984)

Annotation: SIGN observational studies filter

24	7 and 23 (9720)

Annotation: cannabis AND observational studies

25	randomized controlled trial/ (597702)

26	Controlled clinical study/ (463832)

27	random$.ti,ab. (1518977)

28	randomization/ (86491)

29	intermethod comparison/ (258334)

30	placebo.ti,ab. (303428)

31	(compare or compared or comparison).ti. (504683)

32	((evaluated or evaluate or evaluating or assessed or assess) and (compare or compared or comparing or comparison)).ab. (2082229)

33	(open adj label).ti,ab. (78190)

34	((double or single or doubly or singly) adj (blind or blinded or blindly)).ti,ab. (229917)

35	double blind procedure/ (171048)

36	parallel group$1.ti,ab. (25201)

37	(crossover or cross over).ti,ab. (104010)

38	((assign$ or match or matched or allocation) adj5 (alternate or group$1 or intervention$1 or patient$1 or subject$1 or participant$1)).ti,ab. (325625)

39	(assigned or allocated).ti,ab. (383429)

40	(controlled adj7 (study or design or trial)).ti,ab. (343515)

41	(volunteer or volunteers).ti,ab. (244577)

42	human experiment/ (490389)

43	trial.ti. (295850)

44	or/25-43 (4952112)

Annotation: Cochrane RCT filter

45	7 and 44 (14036)

Annotation: cannabis AND RCTs

46	24 or 45 (21357)

Annotation: cannabis AND (Obs studies OR RCTs)

47	7 and (23 or 44) (21357)

Annotation: logic check

48	(ae or si or to or co).fs. (3204803)

49	(safe or safety).ti,ab. (1154971)

50	side effect$.ti,ab. (358075)

51	((adverse or undesirable or harm$ or serious or toxic) adj3 (effect$ or reaction$ or event$ or outcome$)).ti,ab. (787739)

52	exp adverse drug reaction/ (522775)

53	exp drug toxicity/ (125051)

54	exp intoxication/ (366563)

55	exp drug safety/ (393912)

56	exp drug monitoring/ (53058)

57	exp drug hypersensitivity/ (56248)

58	exp postmarketing surveillance/ (35831)

59	exp drug surveillance program/ (26017)

60	exp phase iv clinical trial/ (3822)

61	(toxicity or complication$ or noxious or tolerability).ti,ab. (1868476)

62	or/48-61 (6002309)

Annotation: OVID AE filter 1-14

63	47 and 62 (6382)

Cannabis AEs

Search Name:	cannabis AEs

Date Run:	01/04/2020 18:42:40

Comment:

ID	Search	Hits

#1	MeSH descriptor: [Cannabis] explode all trees	298

#2	MeSH descriptor: [Cannabinoids] explode all trees	790

#3	MeSH descriptor: [Endocannabinoids] explode all trees	48

#4	MeSH descriptor: [Endocannabinoids] explode all trees	48

#5	(Cannabis or cannabinol or cannabinoid* or cannabidiol or bhang or cannador or charas or ganja or ganjah or hashish or hemp or marihuana or marijuana or nabilone or cesamet or cesametic or ajulemic acid or cannabichromene or cannabielsoin or cannabigerol or tetrahydrocannabinol or dronabinol or levonantradol or nabiximols or palmidrol or tetrahydrocannabinolic acid or tetrahydro cannabinol or marinol or tetranabinex or sativex or endocannabinoid*):ti,ab,kw (Word variations have been searched)	4370

#6	#1 or #2 or #3 or #4 or #5	4370

#7	MeSH descriptor: [Drug-Related Side Effects and Adverse Reactions] explode all trees	3463

#8	MeSH descriptor: [] explode all trees and with qualifier(s): [adverse effects - AE, toxicity - TO, poisoning - PO, complications - CO]	169278

#9	(safe or safety):ti,ab,kw (Word variations have been searched)	258304

#10	(side effect*):ti,ab,kw (Word variations have been searched)	149400

#11	((adverse or undesirable or harms* or serious or toxic) near/3 (effect* or reaction* or event* or outcome*)):ti,ab,kw (Word variations have been searched)	279577

#12	MeSH descriptor: [Product Surveillance, Postmarketing] explode all trees	191

#13	MeSH descriptor: [Adverse Drug Reaction Reporting Systems] explode all trees	82

#14	MeSH descriptor: [Clinical Trial, Phase IV] explode all trees	0

#15	MeSH descriptor: [Poisoning] explode all trees	2101

#16	MeSH descriptor: [Substance-Related Disorders] explode all trees	14586

#17	MeSH descriptor: [Abnormalities, Drug-Induced] explode all trees	47

#18	MeSH descriptor: [Drug Monitoring] explode all trees	1725

#19	MeSH descriptor: [Drug Hypersensitivity] explode all trees	965

#20	(toxicity or complication* or noxious or tolerability):ti,ab,kw (Word variations have been searched)	332240

#21	#7 or #8 or #9 or #10 or #11 or #12 or #13 or #14 or #15 or #16 or #17 or #18 or #19 or #20	626064

#22	#6 and #21	in Trials	2426

PsycInfo

Database: APA PsycInfo <1806 to March Week 4 2020>

Search Strategy:

--------------------------------------------------------------------------------

1	exp cannabis/ or exp cannabinoids/ or tetrahydrocannabinol/ (12819)

2	(Cannabis or cannabinol or cannabinoid* or cannabidiol or bhang or cannador or charas or ganja or ganjah or hashish or hemp or marihuana or marijuana or nabilone or cesamet or cesametic or ajulemic acid or cannabichromene or cannabielsoin or cannabigerol or tetrahydrocannabinol or dronabinol or levonantradol or nabiximols or palmidrol or tetrahydrocannabinolic acid or tetrahydro cannabinol or marinol or tetranabinex or sativex or endocannabinoid*).mp. [mp=title, abstract, heading word, table of contents, key concepts, original title, tests & measures, mesh] (26466)

3	1 or 2 (26466)

4	exp "side effects (drug)"/ (57604)

5	(safe or safety).ti,ab. (84148)

6	side effect$.ti,ab. (31950)

7	((adverse or undesirable or harms$ or serious or toxic) adj3 (effect$ or reaction$ or event$ or outcome$)).ti,ab. (44183)

8	toxic disorders/ (1433)

9	exp "substance use disorder"/ (127742)

10	(toxicity or complication$ or noxious or tolerability).ti,ab. (42844)

11	or/4-10 (310848)

12	3 and 11 (10984)

13	epidemiology/ (49562)

14	((case* adj5 control*) or (case adj3 comparison*) or case-comparison or control group*).ti,ab,id. not "Literature Review".md. (95810)

15	((cohort or longitudinal or prospective or retrospective).ti,ab,id. or longitudinal study.md. or prospective study.md. or retrospective study.md.) not "Literature Review".md. (286455)

16	(cross section* or "prevalence study").ti,ab,id. (80384)

17	clinical trials/ or "treatment outcome clinical trial".md. or ((randomi?ed adj7 trial*) or ((single or doubl* or tripl* or treb*) and (blind* or mask*)) or (controlled adj3 trial*) or (clinical adj2 trial*)).ti,ab,id. (101001)

18	Case control.mp. (10736)

19	(cohort adj (study or studies)).mp. [mp=title, abstract, heading word, table of contents, key concepts, original title, tests & measures, mesh] (21026)

20	Cohort analy$.mp. (2099)

21	(Follow up adj (study or studies)).mp. [mp=title, abstract, heading word, table of contents, key concepts, original title, tests & measures, mesh] (12876)

22	(Longitudinal or Retrospective or Cross sectional).mp. [mp=title, abstract, heading word, table of contents, key concepts, original title, tests & measures, mesh] (218589)

23	or/13-22 (561443)

24	12 and 23 (3801)

### Appendix 2: Detailed methods for the assessment of risk of bias

We rated studies at serious risk of confounding bias when they when they did not adjust for important predictors of adverse events and cannabis use, including, at minimum, pain intensity, concomitant pain medication, disability status, alcohol use, past cannabis use and at critical risk if they did not include a control group. We rated studies at serious risk of selection bias when studies included prevalent medical cannabis users (i.e., patients who experience serious or debilitating adverse events are more likely to discontinue cannabis and hence less likely to be included in studies of prevalent users). We rated studies at serious risk of misclassification of the intervention if there was evidence that medical cannabis users were not appropriately classified. We rated studies at serious risk of bias due to departure from the intended intervention if the intervention was not delivered as intended or more than 20% of patients discontinued the intervention for reasons unrelated to adverse effects (e.g., costs). We rated studies at serious risk of missing data when 20% or more of the original patients did not have adverse event data. Finally, we rated studies at moderate risk of selective reporting when the study did not differentiate between minor and serious adverse events or when there were indications that adverse events were selectively, and not comprehensively, reported.

### Appendix 3: List of included studies

1.	Anderson SP, Zylla DM, McGriff DM, Arneson TJ. Impact of medical cannabis on patient-reported symptoms for patients with cancer enrolled in Minnesota's medical cannabis program. Journal of Oncology Practice. 2019;15(6):E338-E45.

2.	Bellnier T, Brown GW, Ortega TR. Preliminary evaluation of the efficacy, safety, and costs associated with the treatment of chronic pain with medical cannabis. The Mental Health Clinician. 2018;8(3):110-5.

3.	Bestard JA, Toth CC. An open-label comparison of nabilone and gabapentin as adjuvant therapy or monotherapy in the management of neuropathic pain in patients with peripheral neuropathy. Pain Practice. 2011;11(4):353-68.

4.	Bonar EE, Cranford JA, Arterberry BJ, Walton MA, Bohnert KM, Ilgen MA. Driving under the influence of cannabis among medical cannabis patients with chronic pain. Drug & Alcohol Dependence. 2019;195:193-7.

5.	Cervigni M, Nasta L, Schievano C, Lampropoulou N, Ostardo E. Micronized Palmitoylethanolamide-Polydatin Reduces the Painful Symptomatology in Patients with Interstitial Cystitis/Bladder Pain Syndrome. BioMed Research International. 2019;2019 (no pagination)(9828397).

### Appendix 4: Studies excluded at the full-text screening stage

**Not a full-text report of a non-randomized study**

12.	Arboleda MF, Dam V, Prosk E, Dworkind M, Vigano A. Tranforming symptom management in cancer patients: Is medical cannabis a new paradigm? Supportive Care in Cancer. 2018;26 (2 Supplement 1):S53.

13.	Ashton CH. Adverse effects of cannabis and cannabinoids. British Journal of Anaesthesia. 1999;83(4):637-49.

14.	Ballas SK. Use of marijuana in patients with sickle cell disease increased the frequency of hospitalization for acute painful vaso-occlusive crises. Blood Conference: 58th Annual Meeting of the American Society of Hematology, ASH. 2016;128(22).

15.	Bergamaschi V, Konrad G, Battaglia MA, Brichetto G. Efficacy and discontinuation of nabiximols in patients with multiple sclerosis: A real-life study. Multiple Sclerosis Journal. 2018;24 (2 Supplement):959.

16.	Bertsche T, Schulz M. Cannabis can relieve spasticity associated with multiple sclerosis. [German]. Pharmazeutische Zeitung. 2003;148(8):32-3.

17.	Bialas P, Drescher B, Gottschling S, Juckenhofel S, Konietzke D, Kuntz W, et al. [Cannabis-based medicines for chronic pain: indications, selection of drugs, effectiveness and safety : Experiences of pain physicians in Saarland]. Der Schmerz. 2019;33(5):399-406.

18.	Blondin N. The evolving role of complementary cannabis therapy in glioblastoma treatment. Neuro-Oncology. 2018;20 (Supplement 6):vi214-vi5.

19.	Bronstein K, Dhaliwal J, Leider H. Rates of inappropriate drug use in the chronic pain population: An update. Journal of Pain. 2011;1):P5.

20.	Brusberg M, Kang D, Larsson H, Lindstrom E, Martinez V. Inhibition of fatty acid amide hydrolase (FAAH) activity enhances the analgesic action of the endocannabinoid anandamide on visceral pain. Gastroenterology. 2009;1):A141.

21.	Bulbul A, Mino EA, Khorsand-Sahbaie M, Lentkowski L. Opioid dose reduction and pain control with medical cannabis. Journal of Clinical Oncology Conference. 2018;36(34 Supplement).

30.	Dimou T, Spanomanoli A, Michelis S. The use of palmitoylethinolamide (PEA) in FBSS for chronic pain management. Regional Anesthesia and Pain Medicine. 2019;44 (10 Supplement 1):A168.

31.	Donato F, Turri M, Zanette G, Tugnoli V, Deotto L, Teatini F, et al. A study of cortical and spinal excitability in patients affected by multiple sclerosis and spasticity after oromucosal cannabinoid spray (THC/CBD). Clinical Neurophysiology. 2016;127 (4):e147.

32.	Donovan KA. Age-related differences in cannabis use by cancer patients referred for supportive care. Diane Portman. Journal of Clinical Oncology Conference. 2019;37(31 Supplement 1).

33.	Dow GJ, Meyers FH, Stanton W, Devine ML. Serious reactions to oral delta-9-tetrahydrocannabinol in cancer chemotherapy patients. Clinical Pharmacy. 1984;3(1):14.

34.	Dusi V, Attili SVS, Singaraju M. Observational study on role of crude cannabis in pain control and quality of life in terminally ill cancer patients: An Indian perspective. Annals of Oncology. 2019;30 (Supplement 9):ix119.

35.	Eltayb A, Etges T, Wright S. An observational post-approval registry study of patients prescribed Sativex. Results from clinical practice. Multiple Sclerosis. 2013;1):480.

48.	Ferrante F, Polito G, Ferraro M. DELTA-9-tetrahydrocannabinol (Sativex) for the treatment of multiple sclerosis spasticity: Evaluation of effectiveness and safety. European Journal of Hospital Pharmacy. 2019;26 (Supplement 1):A239.

49.	Ferre L, Nuara A, Pavan G, Radaelli M, Moiola L, Rodegher M, et al. Medium and long term efficacy of nabiximols for the treatment of multiple sclerosis related spasticity: An Italian monocentric study. Multiple Sclerosis. 2015;1):728-9.

50.	Ferre L, Pavan G, Nuara A, Radaelli M, Liberatore G, Guaschino C, et al. Efficacy, safety and response rate to Nabiximol for the treatment of MS-related spasticity in an Italian monocentric cohort. Multiple Sclerosis. 2015;21 (4):501-2.

51.	Ferre L, Sorosina M, Santoro S, Moiola L, Rodegher M, Colombo B, et al. Efficacy, safety and response rate of nabiximols assessed in an Italian monocentric cohort. Multiple Sclerosis. 2014;1):469-70.

52.	Fitzcharles MA, McDougall J, Ste-Marie PA, Padjen I. Clinical implications for cannabinoid use in the rheumatic diseases: potential for help or harm? Arthritis & Rheumatism. 2012;64(8):2417-25.

53.	Flachenecker P, Zettl U, Henze T. THC:CBD oromucosal spray (nabiximols) in the long term treatment of multiple sclerosis spasticity. The MOVE 2 long-term study. Multiple Sclerosis. 2013;1):527.

59.	Galvin D, Mulkerrin O. Cannabis-based medications: A comparison of patients' knowledge and awareness in pain, neurology and prescription out-patient settings. Pain Practice. 2018;18 (Supplement 1):60.

60.	Gamaoun R, Kasvis P, Patronidis F, Arboleda MF, Vigano A. Potential impact of medical cannabis treatment on pain control among cancer patients in Quebec-Canada: A pilot study. Supportive Care in Cancer. 2019;27 (1 Supplement):S54-S5.

61.	Gaston T, Szaflarski M, Hansen B, Grayson L, Bebin EM, Szaflarski J. Improvement in quality of life ratings after one year of treatment with pharmaceutical formulation of cannabidiol (CBD). Epilepsia. 2017;58 (Supplement 5):S159.

62.	Gauter B, Rukwied R, Konrad C. [Use and effectiveness of dronabinol (delta9-tetrahydrocannabinol) in chronic pain]. Der Schmerz. 2004;18 Suppl 2:S11-4.

63.	Gerardi MC, Batticciotto A, Talotta R, Ditto MC, Atzeni F, Sarzi-Puttini P. Efficacy of cannabis flos in patients with fibromyalgia: A monocentric observational study. Arthritis and Rheumatology. 2016;68 (Supplement 10):72-4.

64.	Gilmore D, Hooper C, Nemastil CJ, Dell ML, McCoy K, Kirkby SE. Effects of self-reported marijuana use on adherence and mental health disease in cystic fibrosis. Pediatric Pulmonology. 2018;53 (Supplement 2):424.

65.	Gubbiotti M, Illiano E, Costantini E, Giannantoni A. Palmitoylethanolamide/polydatin as add-on therapy in pain resistant patients with interstitial cystitis/bladder painful syndrome. European Urology, Supplements. 2019;18 (1):e1970.

66.	Guerrero LJ, MacIas IC, Del Castillo SSF, Izquierdo MM, Rengifo CD, Nunez MN. Effectiveness and safety of d-9-tetrahydrocannabinol (sativex) in patients with multiple sclerosis spasticity. European Journal of Hospital Pharmacy. 2017;24 (Supplement 1):A113.

67.	Guindon J. Nabilone in inflammatory pain: to be or not to be. Clinical & Experimental Pharmacology & Physiology. 2012;39(4):327-8.

68.	Gurevich T, Bar Lev Chleider L, Rosenberg A, Knaani J, Baruch Y, Djaldetti R. Effect of medical cannabis in Parkinson's disease: Survey of patient experiences. Movement Disorders. 2015;1):S88-S9.

69.	Gutierrez T, Hohmann AG. Cannabinoids for the treatment of neuropathic pain: Are they safe and effective? Future Neurology. 2011;6(2):129-33.

70.	Guttenthaler V, Wittmann M. Replacement of benzodiacepines by cannabinoids for the preoperative medication-a feasibility trial (Beach-Trial). Medical Cannabis and Cannabinoids. 2019;2 (2):78.

71.	Gyang T, Hyland M, Samkoff L, Goodman A. "Real world" experience of medical marijuana in symptomatic management of multiple sclerosis and transverse myelitis. Neurology Conference: 70th Annual Meeting of the American Academy of Neurology, AAN. 2018;90(15 Supplement 1).

72.	Hansra D, Granada H. Evaluation of safety, efficacy, and clinical endpoints of delta-9-tetrahydrocannabinol in patients age 60 or older with hematologic and oncologic malignancies. Blood Conference: 59th Annual Meeting of the American Society of Hematology, ASH. 2017;130(Supplement 1).

73.	Hansra DM. Evaluation of safety, efficacy, and other clinical endpoints of delta-9-tetrahydrocannabinol in older patients with hem/onc malignancies. Journal of Clinical Oncology Conference. 2017;35(15 Supplement 1).

74.	Haupts M, Jonas A, Witte K, Alvarez-Ossorio L. Influence of optimized anti-spastic pre-treatment on the efficacy and tolerability of THC: CBD oromucosal spray in multiple sclerosis spasticity patients. A post-hoc RCT data analyses. Multiple Sclerosis. 2015;1):708-9.

75.	Hicks K, Snyder C. Impact of high-dose cannabis use in patients with advanced pancreatic cancer undergoing treatment in a phase i clinical trial: Lessons learned and impact on future clinical research design. Journal of Oncology Pharmacy Practice. 2018;24 (2 Supplement 1):8.

76.	Higgins P, Ginsburg D, Gilder K, Walsh B, English B, Turner S, et al. Safety and efficacy of olorinab, a peripherally restricted, highly-selective, cannabinoid receptor 2 agonist in a phase 2A study in chronic abdominal pain associated with Crohn's disease. Journal of Crohn's and Colitis. 2019;13 (Supplement 1):S318.

77.	Hill KP, Hurley-Welljams-Dorof WM. Low to moderate quality evidence demonstrates the potential benefits and adverse events of cannabinoids for certain medical indications. Evidence Based Medicine. 2016;21(1):17.

78.	Hobart JC, Zajicek JP. Cannabis as a symptomatic treatment for MS: Clinically meaningful MUSEC to the stiffness and walking problems of people with MS. Multiple Sclerosis. 2012;1):247.

79.	Hoffenberg E, Murphy B, Mikulich-Gilbertson S, McWilliams S, Hoffenberg A, Hopfer C. Why and how adolescents and young adults with inflammatory bowel disease use cannabis. Journal of Pediatric Gastroenterology and Nutrition. 2017;65 (Supplement 2):S147-S8.

80.	Honarmand K, Tierney MC, O'Connor P, Feinstein A. Effects of cannabis on cognitive function in patients with multiple sclerosis. Neurology. 2011;76(13):1153-60.

81.	Huestis MA, Elsohly M, Nebro W, Barnes A, Gustafson RA, Smith ML. Estimating time of last oral ingestion of cannabis from plasma THC and THCCOOH concentrations. Therapeutic Drug Monitoring. 2006;28(4):540-4.

82.	Hulgan T, Kingsley P, Koethe J, Sterling T, Patel S. Associations between circulating endocannabinoids and cardio-metabolic factors in HIV-infected persons on antiretroviral therapy: A pilot study. Antiviral Therapy. 2014;2):A8.

89.	Khalid L, Starrels JL, Sohler N, Arnsten JH, Jost J, Cunningham C. Marijuana use is associated with low prescription opioid analgesic (POA) use among hiv-infected patients with chronic pain. Journal of General Internal Medicine. 2016;1):S297.

90.	Kiszko K, Patel K, Chudasama B, Samodulski J, Nienaber C, Martins-Welch D, et al. Older adults' perspectives on medical marijuana (MM) use. Journal of the American Geriatrics Society. 2017;65 (Supplement 1):S70.

91.	Klooker T, Leliefeld K, Van Den Wijngaard RM, Boeckxstaens GE. The cannabinoid receptor agonist delta-9-tetrahydrocannabinol increases rectal sensitivity in IBS patients and healthy volunteers. Gastroenterology. 2009;1):A726-A7.

92.	Koehler J, Feneberg W, Gorodetzky H, Meier M, Pollmann W. Clinical experiences with on-label nabiximols therapy in multiple sclerosis-induced spasticity. Multiple Sclerosis. 2013;1):281-2.

93.	Koehler J, Gorodetzky H, Pollmann W, Meier M, Feneberg W. Monotherapy with nabiximols in multiple sclerosisinduced spasticity. Multiple Sclerosis. 2013;1):282.

102.	Macari D, Gbadamosi B, Ezekwudo D, Khoury J, Jaiyesimi IA, Gaikazian SS. Medical cannabis in cancer patients: Prevalence, efficacy, and safety. Journal of Clinical Oncology Conference. 2019;37(Supplement 15).

103.	Maggioli C, Giannone FA, Baldassarre M, Fanelli F, Mezzullo M, Belluomo I, et al. Endocannabinoids in advanced cirrhosis: Have we picked the right one? Digestive and Liver Disease. 2012;1):S40.

104.	Malfitano AM, Laezza C, D'Alessandro A, Procaccini C, Saccomanni G, Tuccinardi T, et al. Effects on immune cells of a new 1,8-naphthyridin-2-one derivative and its analogues as selective CB2 agonists: implications in multiple sclerosis. PLoS ONE [Electronic Resource]. 2013;8(5):e62511.

105.	Martellucci I, Laera L, Lippi S, Marsili S, Petrioli R, Francini G. Impact of cannabinoids on the quality of life in oncology: Prospective observational study. Annals of Oncology Conference: 17th National Congress of Medical Oncology Rome Italy Conference Publication:. 2015;26(SUPPL. 6).

106.	Mbachi C, Wang Y, Barkin JA, Demetria MV, Barkin JS, Kroner PT, et al. Does cannabis consumption impact chronic pancreatitis related complications? American Journal of Gastroenterology. 2019;114 (Supplement):S21-S2.

107.	Mc Vige J, Bargnes VH, Shukri S, Mechtler L. Cannabis, concussion, and chronic pain: An ongoing retrospective analysis at Dent Neurologic Institute in Buffalo, NY. Neurology. 2018;91 (23 Supplement 1):S18-S9.

108.	McLeod SA, Lemay JF. Medical cannabinoids. CMAJ Canadian Medical Association Journal. 2017;189(30):E995.

109.	McQuay HJ. More evidence cannabis can help in neuropathic pain. CMAJ Canadian Medical Association Journal. 2010;182(14):1494-5.

110.	McVige J, Kaur D, Hart P, Lillis M, Mechtler L, Bargnes V, et al. Medical cannabis in the treatment of post-traumatic concussion. Neurology Conference: 71st Annual Meeting of the American Academy of Neurology, AAN. 2019;92(15 Supplement 1).

111.	Mechtler L, Bargnes V, Hart P, McVige J, Saikali N. Medical cannabis for chronic migraine: A retrospective review. Neurology Conference: 71st Annual Meeting of the American Academy of Neurology, AAN. 2019;92(15 Supplement 1).

112.	Mechtler L, Hart P, Bargnes V, Saikali N. Medical cannabis treatment in patients with trigeminal neuralgia. Neurology Conference: 71st Annual Meeting of the American Academy of Neurology, AAN. 2019;92(15 Supplement 1).

122.	Morrison G, Sardu ML, Rasmussen CH, Sommerville K, Roberts C, Blakey GE. Exposure-response analysis of cannabidiol for the treatment of lennox-gastaut syndrome. Epilepsia. 2018;59 (Supplement 3):S11-S2.

123.	Morrison G, Sardu ML, Rasmussen CH, Sommerville K, Roberts C, Blakey GE. Exposure-Response Analysis of Cannabidiol (CBD) oral solution for the treatment of lennox-gastaut syndrome. Neurology Conference: 70th Annual Meeting of the American Academy of Neurology, AAN. 2018;90(15 Supplement 1).

124.	Mousa A, Petrovic M, Laszlo S, Fleshner N. Is there a therapeutic role for cannabis in advanced prostate cancer? Exploring the patterns and predictors of use among men receiving androgen-deprivation therapy. Canadian Urological Association Journal. 2018;12 (6 Supplement 2):S126.

125.	Mupamombe CT, Nathan RA, Case AA, Walter M, Hansen E. Efficacy of medical cannabis for cancer-related pain in the elderly: A single-center retrospective analysis. Journal of Clinical Oncology Conference. 2019;37(31 Supplement 1).

126.	Myers B, Geist T, Hart P, Aladeen T, Begley A, Westphal ES, et al. Medical cannabis in the treatment of parkinson's disease. Neurology Conference: 71st Annual Meeting of the American Academy of Neurology, AAN. 2019;92(15 Supplement 1).

127.	Nadal X, Del Rio C, Casano S, Palomares B, Ferreiro-Vera C, Navarrete C, et al. Tetrahydrocannabinolic acid is a potent PPARgamma agonist with neuroprotective activity. British Journal of Pharmacology. 2017;174(23):4263-76.

128.	Naftali T, Bar Lev Schlieder L, Hirsch J, Lish I, Benjaminov F, Konikoff F. Cannabis use patterns in patients with IBD. Journal of Crohn's and Colitis. 2016;10 (Supplement 1):S375-S6.

129.	Naftali T, Bar Lev Schlieder L, Sklerovsky Benjaminov F, Lish I, Hirsch J, Konikoff FM. Cannabis induces clinical and endoscopic improvement in moderately active ulcerative colitis (UC). Journal of Crohn's and Colitis. 2018;12 (Supplement 1):S306.

130.	Naftali T, Bar-Lev L, Gabay G, Chowers Y, Dotan I, Bronshtein M, et al. Tetrahydrocannabinol (THC) rich medical cannabis induces clinical and biochemical improvement with a steroid sparing effect in active crohn's disease. Gastroenterology. 2012;1):S780.

131.	Naftali T, Barlev L, Gabay G, Chowers Y, Dotan I, Stein A, et al. Tetrahydrocannabinol (THC) induces clinical and biochemical improvement with a steroid sparing effect in active inflammatory bowel disease. Journal of Crohn's and Colitis. 2013;7 (SUPPL.1):S153.

132.	Nathan RA, Tonderai C, Mupamombe, Walter M, Case AA, Hansen E. Use of medical cannabis in treating anorexia and nausea in elderly cancer patients. Journal of Clinical Oncology Conference. 2019;37(31 Supplement 1).

153.	Nct. Phase 1 Study to Study the Efficacy and Safety of Cannabis in the Treatment of Tinnitus. Clinicaltrialsgov [wwwclinicaltrialsgov]. 2013.

154.	Nct. Safety and Efficacy of Nabilone in Alzheimer's Disease. http0s://clinicaltrialsgov/show/NCT02351882. 2014.

177.	Ngan TYT, Litt M, Eguzo K, Thiel JA. Patient Outcomes Following Initiation of Medical Cannabis in Women with Chronic Pelvic Pain. Journal of Minimally Invasive Gynecology. 2019;26 (7 Supplement):S89-S90.

178.	Nickels K. Cannabidiol in patients with intractable epilepsy due to TSC: A possible medication but not a miracle. Epilepsy Currents. 2017;17(2):91-2.

179.	Nicolodi M, Pinnaro MS, Sandoval V. Selected cannabinoids and cutaneous allodynia in chronic refractory migraine. Journal of Headache and Pain Conference: 12th European Headache Federation Congress and the 32nd National Congress of the Italian Society for the Study of Headaches Italy. 2018;19(Supplement 1).

180.	Nicolodi M, Pinnaro MS, Sandoval V, Torrini A. Possible effects, side-effects and adverse events of a selective cannabinoid (6% THC/7.5% CBD) in refractory chronic migraine. 2013-2018 pilot data. Journal of Headache and Pain Conference: 12th European Headache Federation Congress and the 32nd National Congress of the Italian Society for the Study of Headaches Italy. 2018;19(Supplement 1).

181.	Nicolodi M, Sandoval V, Torrini A. Therapeutic use of cannabinoids-dose finding, effects and pilot data of effects in chronic migraine and cluster headache. European Journal of Neurology. 2017;24 (Supplement 1):287.

182.	Nikles CJ, Yelland M, Glasziou PP, Del Mar C. Do individualized medication effectiveness tests (n-of-1 trials) change clinical decisions about which drugs to use for osteoarthritis and chronic pain? American Journal of Therapeutics. 2005;12(1):92-7.

183.	Notcutt W, Phillips C, Hughes J, Lacoux P, Vijayakulasingam V, Baldock L. A retrospective description of the use of nabilone in UK clinical practice. Multiple Sclerosis. 2014;1):468.

191.	Poli P, Salvadori C, Sannino C. Effects of cannabis based drugs on chronic neuropathic pain: Comparison between italian and dutch medical cannabis variety. Pain Practice. 2018;18 (Supplement 1):101.

192.	Quintans JS, Antoniolli AR, Almeida JR, Santana-Filho VJ, Quintans-Junior LJ. Natural products evaluated in neuropathic pain models - a systematic review. Basic & Clinical Pharmacology & Toxicology. 2014;114(6):442-50.

193.	Reisfield GM. Medical cannabis and chronic opioid therapy. Journal of Pain & Palliative Care Pharmacotherapy. 2010;24(4):356-61.

194.	Reznik I. Post-traumatic stress disorder and medical cannabis use: A naturalistic observational study. European Neuropsychopharmacology. 2012;2):S363-S4.

195.	Reznik I. Medical marijuana/cannabis treatment of Tourette's syndrome: Focus on the quality of life. European Neuropsychopharmacology. 2014;2):S645-S6.

196.	Robinson D, Garti A, Yassin M. Cannabis treatment of diabetic neuropathy: Treatment effect and change in health over a 6 month period. Foot and Ankle Surgery. 2016;1):58.

197.	Ron A, Abuhasira R, Novack V. Establishment of a specialized geriatric clinic providing medical cannabis. Journal of the American Geriatrics Society. 2019;67 (Supplement 1):S299.

198.	Roy A, Konda M, Goel A, Sasapu A. Characteristics of marijuana usage in sickle cell patients. Journal of Investigative Medicine. 2020;68 (2):646.

199.	Roy AM, Konda M, Goel A, Sasapu A. Characteristics of marijuana usage in sickle cell patients: A nationwide analysis. Blood Conference: 61st Annual Meeting of the American Society of Hematology, ASH. 2019;134(Supplement 1).

207.	Schorn M, Krashin D, Mannava A, Belaskova S, Murinova N. Marijuana use in headache in a university-based headache clinic. Neurology Conference: 71st Annual Meeting of the American Academy of Neurology, AAN. 2019;92(15 Supplement 1).

208.	Seibert SM, Kumar P, Gomez PL, Gomez CN, Miller LM, Logsdon M. Cannabis in cancer patients [CP] to improve quality of life [QOL] and cancer related symptoms [CRS]: Illinois cancer care cannabis education and clinical analysis. Journal of Clinical Oncology Conference. 2018;36(15 Supplement 1).

217.	Tripp D, Nickel JC, Laura K, Ginting JV, Mark W, Santor D. Cannabis (marijuana) use in men with chronic prostatitis / chronic pelvic pain syndrome. Journal of Urology. 2012;1):e439-e40.

218.	Trojano M. THC:CBD Observational Study Data: Evolution of Resistant MS Spasticity and Associated Symptoms. European Neurology. 2016;75 Suppl 1:4-8.

219.	Vermersch P, Trojano M. Tetrahydrocannabinol + cannabidiol oromucosal spray for multiple sclerosis resistant spasticity on daily practice, new data. Multiple Sclerosis. 2016;22 (Supplement 3):377.

230.	Wirrell E, Privitera M, Bhathal H, Wong M, Cross J, Sommerville K. Cannabidiol (CBD) treatment effect and adverse events (AES) by time in patients with lennox-gastaut syndrome (LGS): Pooled results from 2 trials. Annals of Neurology. 2018;84 (Supplement 22):S341.

236.	Zettl U, Henze T, Pfiffner C, Vila Silvan C, Flachenecker P. Effectiveness of Sativex in multiple sclerosis spasticity. First data from a large observational study in Germany. Multiple Sclerosis. 2012;1):246.

237.	Zettl UK, Rommer P, Hipp P, Patejdl R. Evidence for the efficacy and effectiveness of THC-CBD oromucosal spray in symptom management of patients with spasticity due to multiple sclerosis. Therapeutic Advances in Neurological Disorders. 2016;9(1):9-30.

238.	Zhou R, Jacobson C, Weng J, Cheng E, Lay J, Hung P, et al. Potential efficacy of cannabidiol for treatment of refractory infantile spasms and lennox gastaut syndrome. Epilepsy Currents. 2015;1):360-1.

239.	Ziemssen T. Tetrahydrocannabinol: Cannabidiol oromucosal spray for treating symptoms of multiple sclerosis spasticity: Newest evidence. Neurodegenerative Disease Management. 2019;9(2s):1-2.

**Study did not include patients with chronic pain**

1.	Abuhasira R, Ron A, Sikorin I, Novack V. Medical cannabis for older patients-treatment protocol and initial results. Journal of Clinical Medicine. 2019;8 (11) (no pagination)(1819).

2.	Adejumo AC, Adegbala OM, Adejumo KL, Bukong TN. Reduced Incidence and Better Liver Disease Outcomes among Chronic HCV Infected Patients Who Consume Cannabis. Canadian Journal of Gastroenterology and Hepatology. 2018;2018 (no pagination)(9430953).

38.	Klotz KA, Grob D, Hirsch M, Metternich B, Schulze-Bonhage A, Jacobs J. Efficacy and Tolerance of Synthetic Cannabidiol for Treatment of Drug Resistant Epilepsy. Frontiers in Neurology. 2019;10 (no pagination)(1313).

**Study did not report on medical cannabis**

**Study did not report on harms or adverse events**

9.	Curtis SA, Spodick J, Lew D, Roberts JD. Medical marijuana certification for patients with sickle cell disease: A survey study of patient's use and preferences. Blood Conference: 60th Annual Meeting of the American Society of Hematology, ASH. 2018;132(Suppl. 1).

19.	Lucas P, Baron EP, Jikomes N. Medical cannabis patterns of use and substitution for opioids & other pharmaceutical drugs, alcohol, tobacco, and illicit substances; Results from a cross-sectional survey of authorized patients. Harm Reduction Journal Vol 16 2019, ArtID 9. 2019;16.

**Study included <25 patients**

1.	Apel A, Greim B, Zettl UK. How frequently do patients with multiple sclerosis use complementary and alternative medicine? Complementary Therapies in Medicine. 2005;13(4):258-63.

2.	Mondello E, Quattrone D, Cardia L, Bova G, Mallamace R, Barbagallo AA, et al. Cannabinoids and spinal cord stimulation for the treatment of failed back surgery syndrome refractory pain. Journal of Pain Research. 2018;11:1761-7.

3.	Toth C, Au S. A prospective identification of neuropathic pain in specific chronic polyneuropathy syndromes and response to pharmacological therapy. Pain. 2008;138(3):657-66.

### Appendix 5: Risk of bias ratings

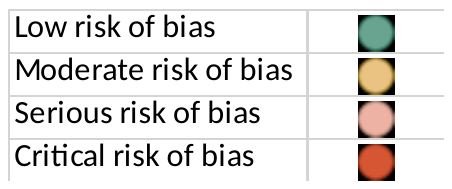

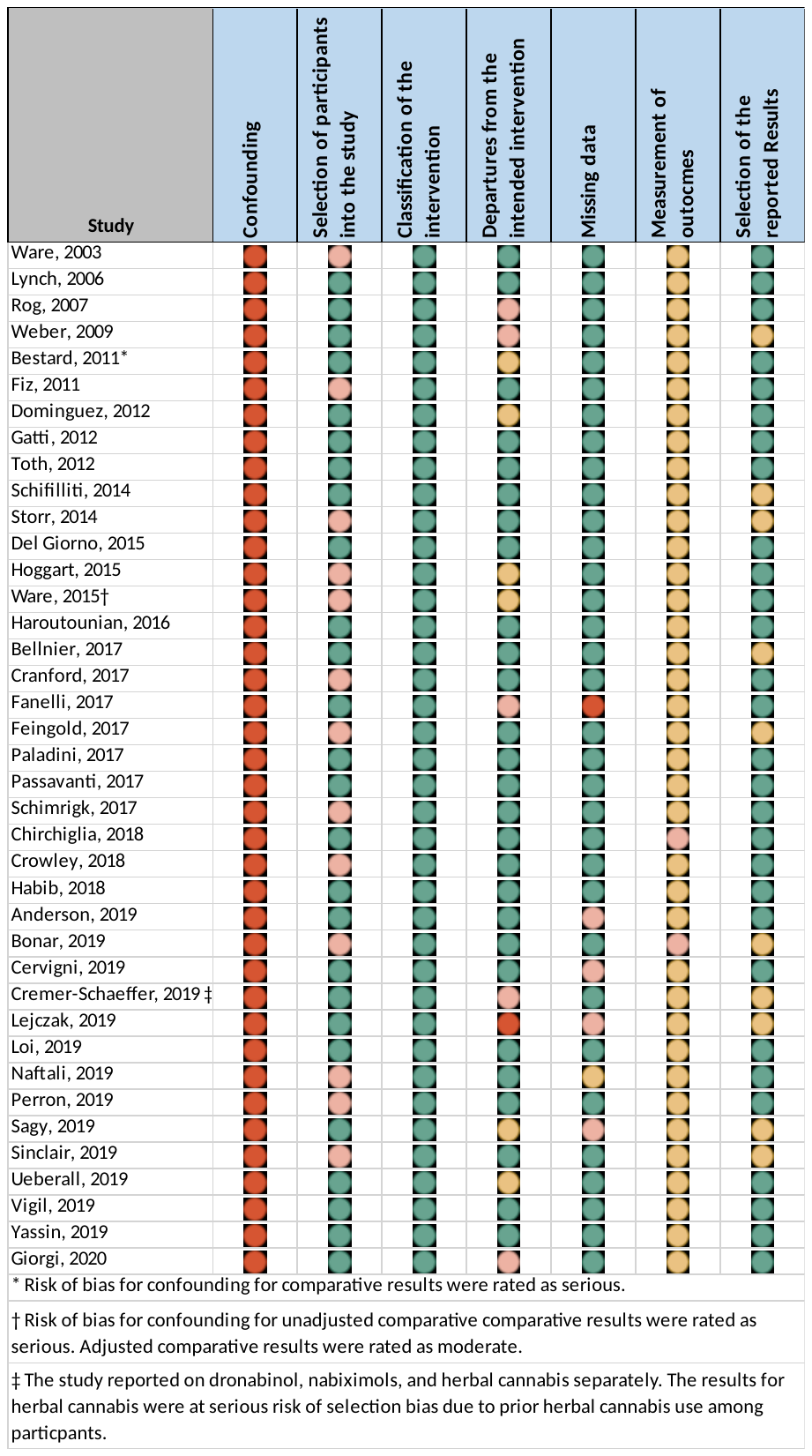

### Appendix 6: Results for all adverse events (subgroup by design)

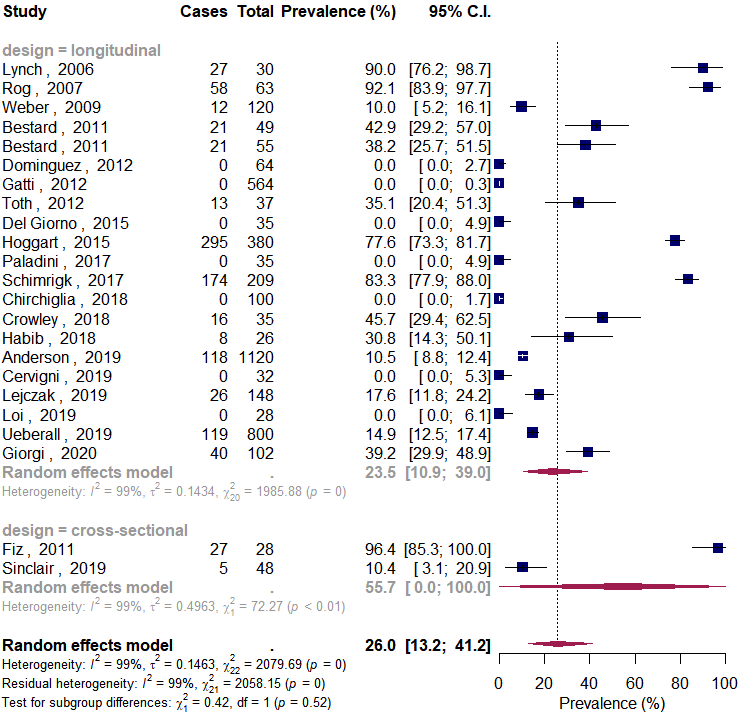

### Appendix 7: Results for all adverse events (subgroup by duration)

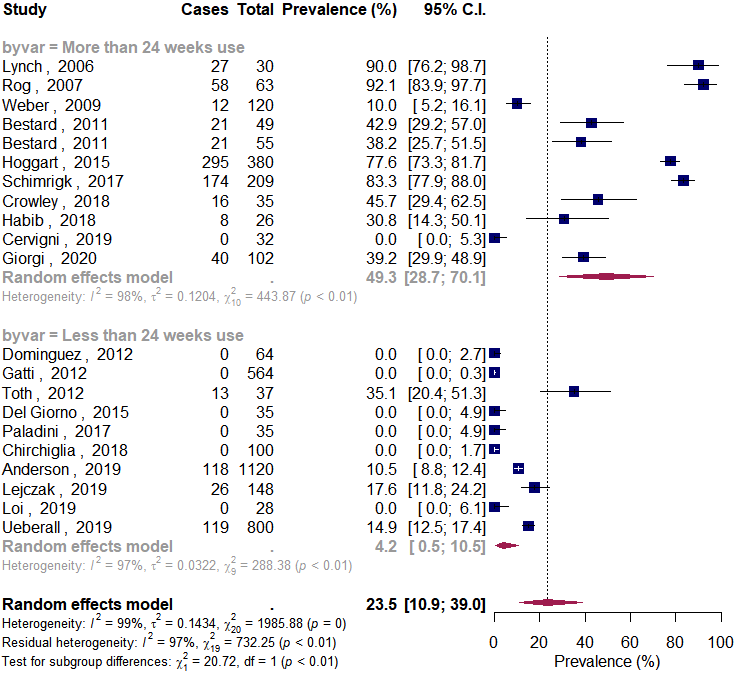

### Appendix 8: Results for all adverse events (subgroup by cannabis)

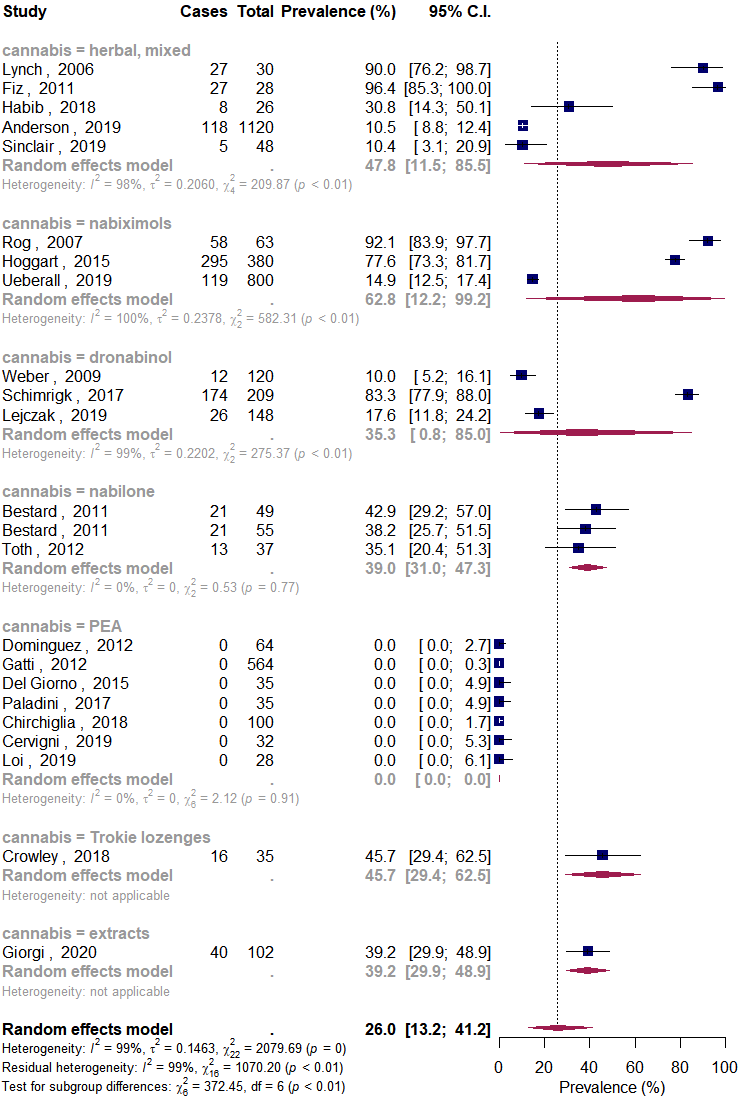

### Appendix 9: Results for all adverse events (subgroup by selection bias)

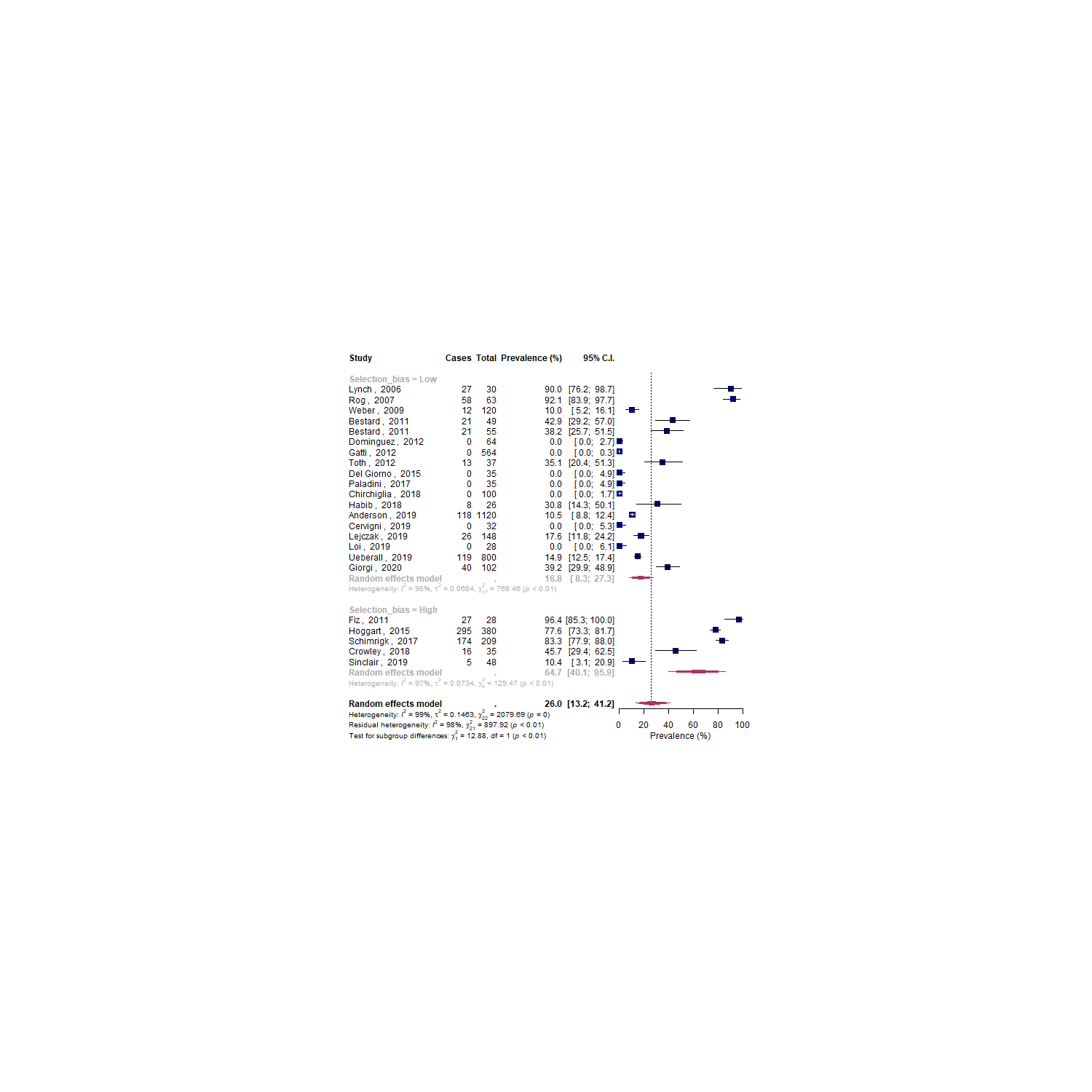

### Appendix 10: Results for adverse events leading to discontinuation (subgroup by duration)

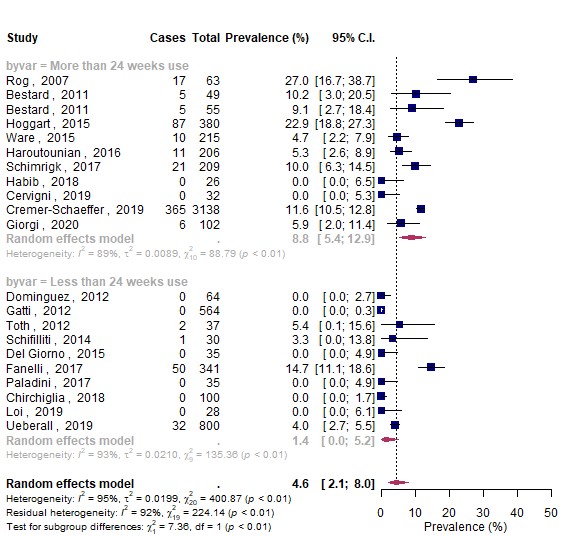

### Appendix 11: Results for adverse events leading to discontinuation (subgroup by cannabis)

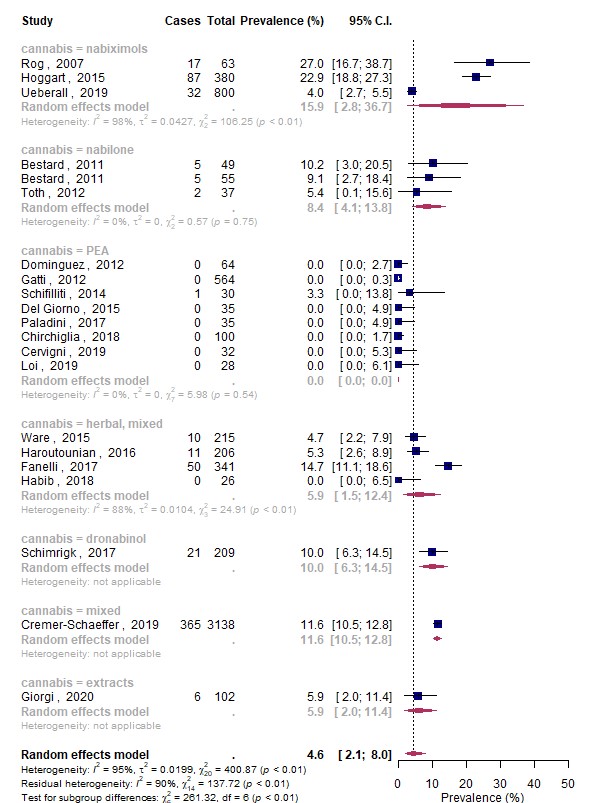

### Appendix 12: Results for adverse events leading to discontinuation (subgroup by selection bias)

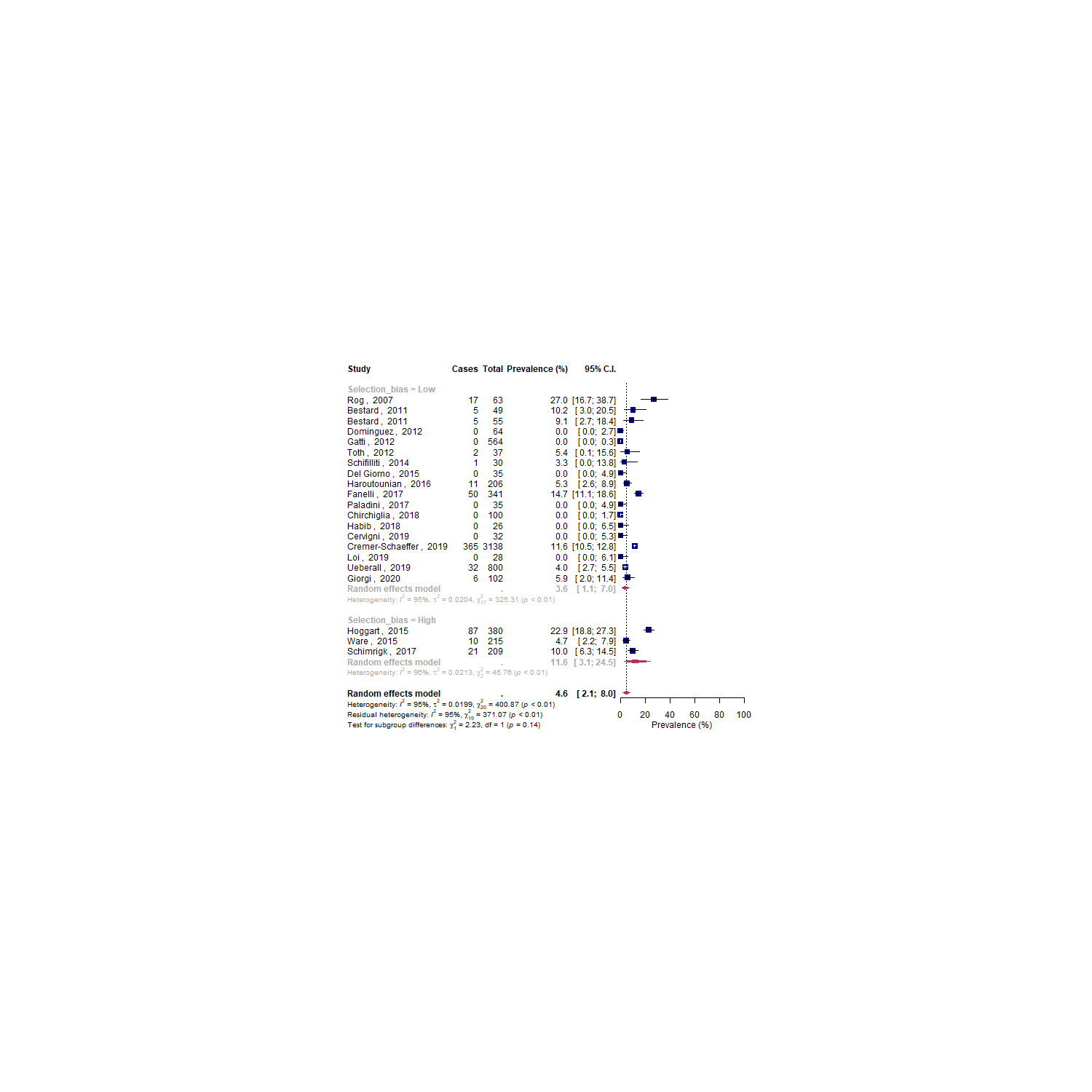

### **Appendix 13: Results for serious adverse events (subgroup by design)**

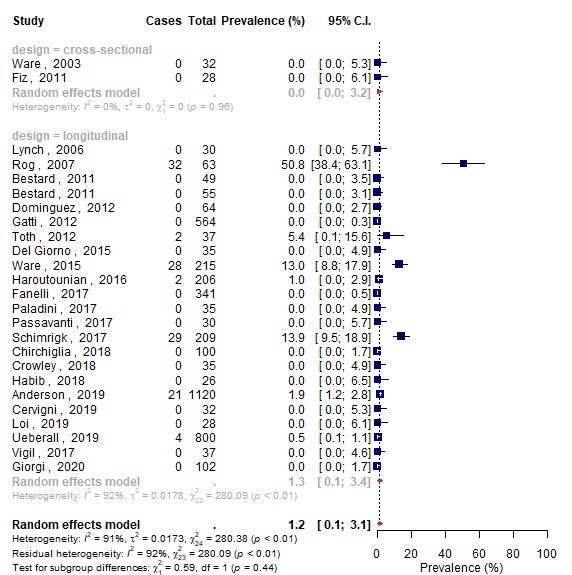

### **Appendix 14: Results for serious adverse events (subgroup by duration)**

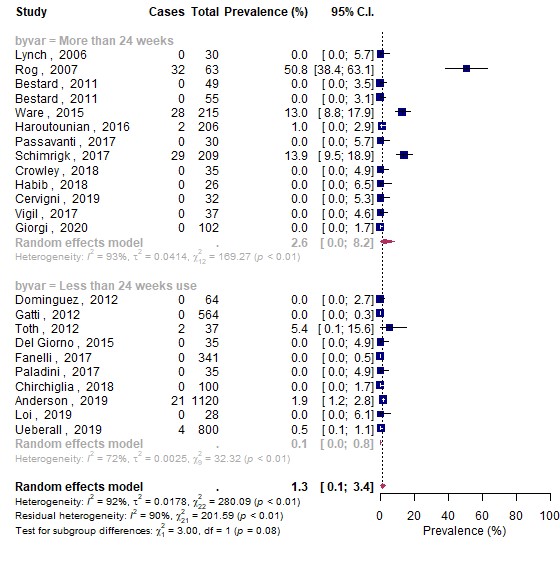

Appendix 15: Results for serious adverse events (subgroup by selection bias)
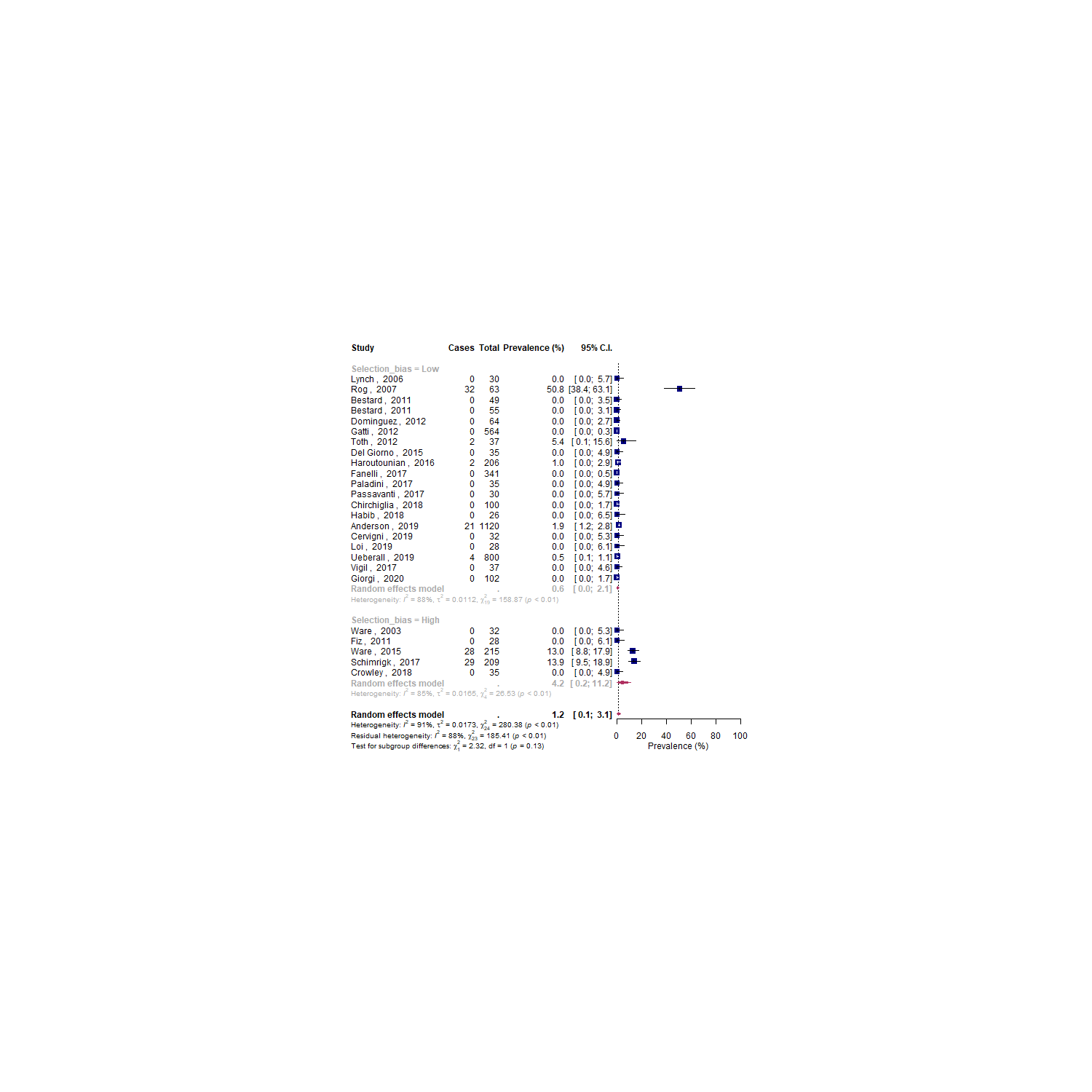

Appendix 16: Results for psychiatric adverse events

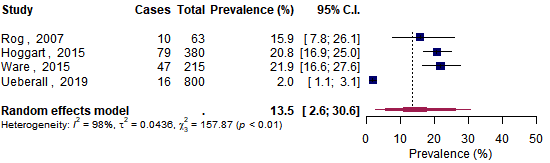

Appendix 17: Results for suicide

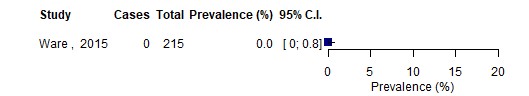

Appendix 18: Results for suicidal thoughts

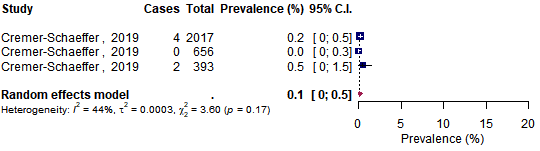

Appendix 19: Results for depression

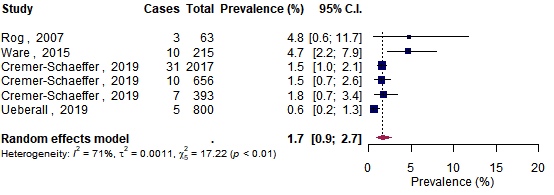

Appendix 20: Results for mania

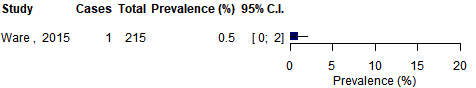

Appendix 21: Results for hallucinations

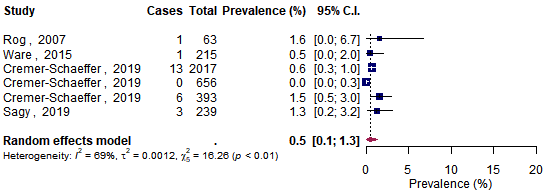

Appendix 22: Results for delusions

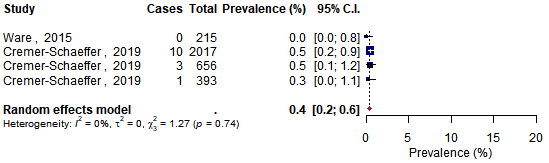

Appendix 23: Results for paranoia

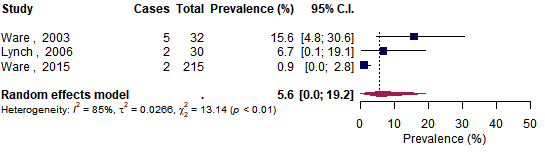

Appendix 24: Results for anxiety

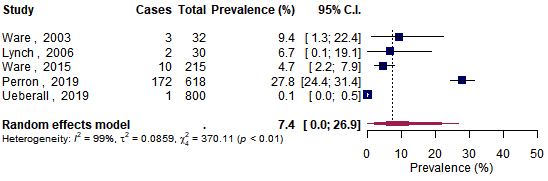

Appendix 25: Results for euphoria

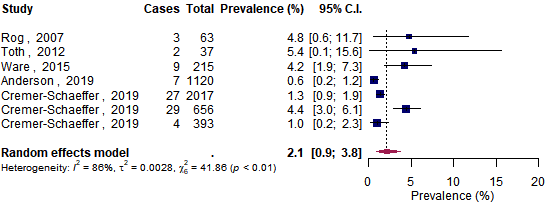

Appendix 26: Results for memory impairment

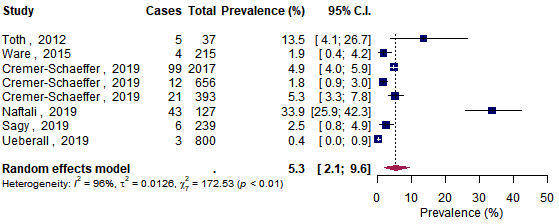

Appendix 27: Results for confusion

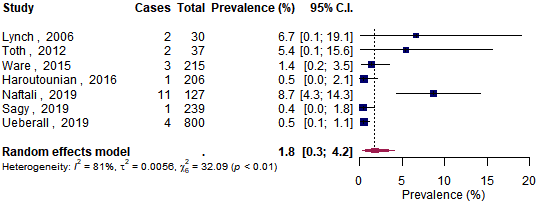

Appendix 28: Results for disorientation

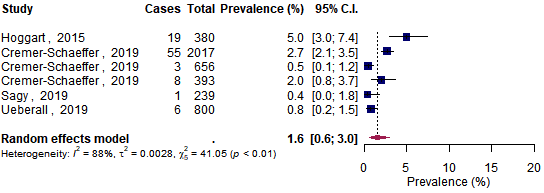

Appendix 29: Results for impaired attention

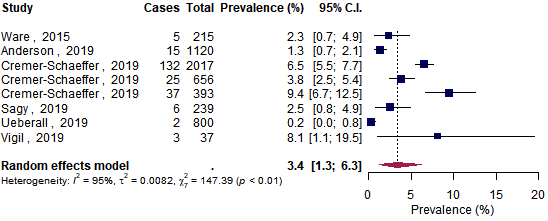

Appendix 30: Results for falls

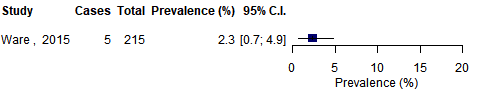

Appendix 31: Results for motor vehicle accidents

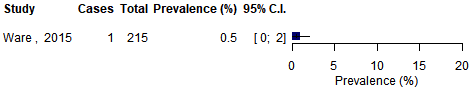

Appendix 32: Results for dependence

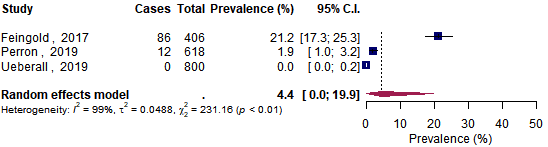

Appendix 33: Results for withdrawal symptoms

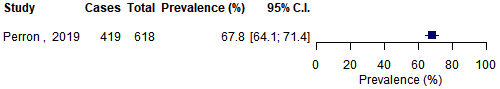

Appendix 34: Results for withdrawal syndrome

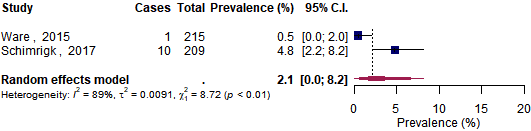
